# Sex-Specific Patterns of Left Ventricular Remodeling Using Regional Wall Thickness Data and Their Associations with Cardiovascular Disease Risk

**DOI:** 10.64898/2026.06.29.26356804

**Authors:** Susanne Rospleszcz, Till Ittermann, Margarethe Woeckel, Sabine Schipf, Christopher Schuppert, Corinna Storz, Roberto Lorbeer, Robin Bülow, Marcus Dörr, Stephan B. Felix, Christian Templin, Henry Völzke, Annette Peters, Fabian Bamberg, Christopher L. Schlett, Marcello Ricardo Paulista Markus

## Abstract

**Background:** Left ventricular (LV) remodeling is associated with impaired cardiac function and future cardiovascular disease (CVD). Current remodeling definitions use broad categorizations based on hypertrophy and mean wall thickness. Cardiac magnetic resonance (CMR) imaging provides detailed characterization of regional myocardial wall properties, which may enhance sex-specific cardiovascular disease (CVD) risk stratification.

**Objectives:** We aimed to identify detailed LV remodeling patterns, their associations with CVD risk, and their clinical predictors.

**Methods:** LV wall thickness data were obtained by CMR in three independent population-based cohorts (SHIP-TREND-0, n=931; SHIP-START-2, n=490; KORA-FF4, n=368). Sex-specific remodeling patterns were identified by k-means clustering and associated with established CVD risk scores and incident morbidity and all-cause mortality. Bootstrapped multinomial regression with LASSO regularization was used to select relevant clinical predictors of remodeling patterns.

**Results:** The sample comprised 991 men (mean age 52.9 years, prevalent CVD 7.8%) and 798 women (52.5 years, 2%). Four remodeling clusters were found for men and women, respectively. For a subset of these clusters, significant associations with an increased CVD risk were found, e.g. in women, the high-risk cluster was associated with a 10.6 (95% confidence interval: 8.9, 12.3) percentage point increase in the 10-year Framingham Risk Score. Associations were independent of blood pressure and myocardial mass. Only in women, associations were also independent of average wall thickness and LV concentricity. Variable selection identified distinct clinical predictors of remodeling patterns.

**Conclusion:** Particularly in women, regional LV wall thickness patterns detect unfavorable cardiac remodeling and might improve CVD risk stratification beyond existing strategies. Automated implementation during image acquisition and integration with shape-based models may facilitate clinical application.

**Graphical Abstract:** 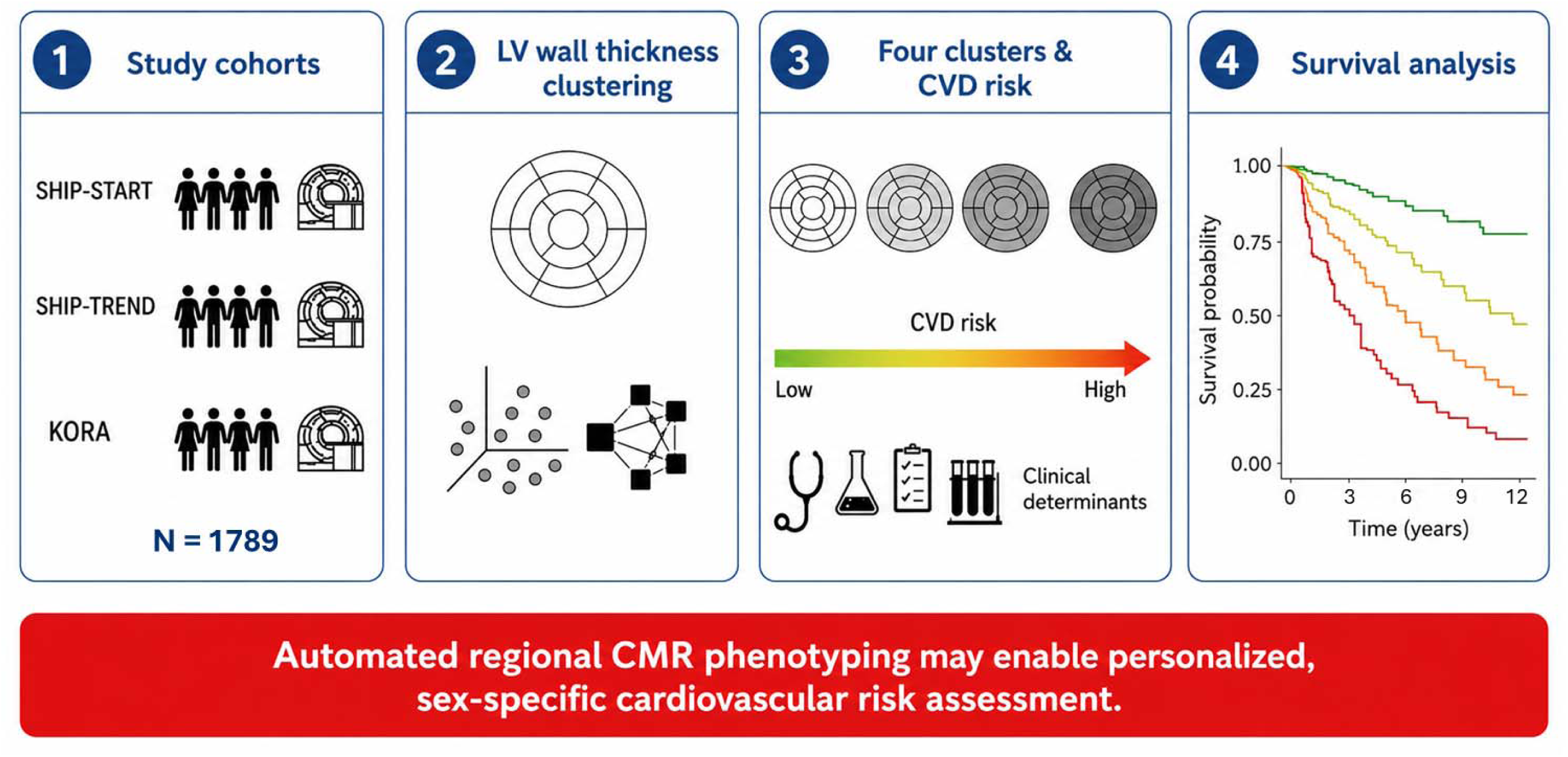

## Introduction

Left ventricular (LV) remodeling is a cardiac adaptation to increased volume or pressure load. Examples are physiological changes due to physical exercise or biological aging, some of which are reversible, adaptive changes after acute myocardial injury, and maladaptive changes due to chronic hemodynamic overload. For the latter, cardiometabolic risk factors such as hypertension [1], obesity and fat distribution [2, 3], hyperglycemia or insulin resistance [4, 5] and impaired renal function [6] are important contributors. Remodeling also represents an intermediate state in CVD development, particularly heart failure. Large cohort studies have shown that LV remodeling is associated with incidence and prevalence of CVD and overall mortality [7, 8] on the population level.

Traditionally, remodeling has been defined by M-mode echocardiography and categorized into either concentric remodeling, eccentric hypertrophy, or concentric hypertrophy [9], but this classification does not account properly for the interaction between wall thickness and chamber dilatation. For this target, a more refined 4-group classification was developed, further dividing the concentric and eccentric categories into two subgroups [10]. Recently a continuous remodeling index based on a biomechanical model of wall stress was designed, incorporating end-diastolic volume and maximum segmental wall thickness [11].

There is an understanding that different patterns of LV morphology provide important information about CVD pathophysiology and risk. However, current classifications are based on prior biomechanical knowledge and preconceptions of regular LV geometry. Agnostic data-driven assessment might provide additional, or even superior, information about individual remodeling patterns that can be used for CVD risk stratification. Radiomic features derived from cardiac magnetic resonance (CMR) imaging of the LV have demonstrated good classification performance in distinguishing different types of cardiac hypertrophy [12]. Recent work from the MESA study showed that LV remodeling signatures based on statistical shape models from CMR are prognostic of CVD at 10-year follow-up [13]. Nonetheless, a large part of the information contained in CMR data is not utilized in current clinical practice.

The development of cardiac remodelling and CVD differs between men and women due to differences in hormonal signalling, metabolism and environmental factors [14]. Both population-and patient-based studies showed that women more often present with concentric hypertrophy and heart failure with preserved ejection fraction, while men more often develop eccentric hypertrophy and heart failure with reduced ejection fraction (EF) [15]. Moreover, the impact of traditional cardiovascular risk factors differs between sexes [16], for example, hypertension and diabetes have been reported to confer higher CVD risk in women [17, 18] and adequate risk factor control might prevent more CVD in women compared to men [19].

Therefore, it is crucial to further explore LV remodeling patterns and their clinical determinants in a sex-specific fashion. In the current study, we aim to utilize CMR data from three independent population-based cohorts to identify sex-specific patterns of LV remodeling and investigate their associations with CVD risk and traditional clinical risk factors.

## Methods

### Study sample

The Study of Health in Pomerania (SHIP) is based on the general adult population of two counties in Northeast Germany [20]. SHIP-START-0, recruited between 1997 and 2001, consists of N=4308 individuals, of whom 2333 participated in the second follow-up SHIP-START-2 between 2008 and 2012. Of these, 1183 participated in a whole-body MRI examination. SHIP-TREND-0, recruited between 2008 and 2012, consists of N=4420 individuals. Of these, 2188 participated in a whole-body MRI examination. Importantly, there is no overlap between participants of both SHIP studies. SHIP was approved by the Ethics Committee of the University of Greifswald and all participants gave written informed consent.

The Cooperative Health Research in the Region of Augsburg (KORA) is based on the general adult population in the region of Augsburg in Southern Germany. KORA-S4, recruited between 1999 and 2001 consists of N=4261 individuals, of whom 2279 participated in the second follow-up KORA-FF4 between 2013 and 2014. Of these, 400 participated in a whole-body MRI examination [21]. KORA was approved by the Bavarian Chamber of Physicians and the Ethics committee of the Ludwig-Maximilians-University Munich and all participants gave written informed consent.

Individuals were excluded from the current study if CMR was not available or if images were of insufficient quality (Figure S1).

### CMR assessment

#### SHIP

The general setup of the whole-body MRI examination has been described previously [20]. In both SHIP-START-2 and SHIP-TREND-0, CMR was performed on 1.5T (MAGNETOM Avanto, Siemens Medical Systems, Erlangen, Germany) with cine steady-state free precession (SSFP) in the short and long axis. In the short axis, the following parameters were used: slice thickness 6mm, FOV 360×292, matrix 256×146, repetition time 2.82ms, echo times 1.2 and 12-14s, flip angle 68°. In the long axis (2 and 4 chamber view), the following parameters were used: slice thickness 6mm, FOV 340×276, matrix 192×125, repetition time 2.65ms, echo time 1.12, flip angle 66°. Late Gadolinium Enhancement (LGE) was determined on a phase-sensitive inversion recovery single-shot SSFP sequence, 15 minutes after intravenous administration of gadobutrol (Gadovist, Bayer Schering Healthcare, Leverkusen, Germany, dosage: 0.15 ml per kg body weight). QMass MR 7.2 (MEDIS, Leiden, Netherlands) was used for semiautomatic image processing. Volume/time curves of the cardiac cycle were obtained by tracing contours of epicardial and endocardial borders at end-diastole and end-systole.

#### KORA

The general setup of the whole-body MRI examination has been described previously [21]. CMR was performed on 3.0T (MAGNETOM Skyra, Siemens Medical Systems, Erlangen, Germany) with SSFP in the short and long axis. In the short axis, the following parameters were used: slice thickness 8mm, FOV 297×360, matrix 240×160, repetition time 29.97ms, echo times 1.46 and 10s, flip angle 62°. In the long axis, the following parameters were used: slice thickness 8mm, FOV 297×360, matrix 240×160, repetition time 29.97ms, echo time 1.46, flip angle 63°. LGE was determined on Fast Low Angle Shot (FLASH) inversion recovery sequences, 10 minutes after intravenous administration of gadopentetate dimeglumine (Gadovist, Bayer Healthcare, Berlin, Germany, dosage: 0.15 ml per kg body weight). cvi42 (Circle Cardiovascular Imaging, Calgary, Canada) was used for semiautomatic image processing. Contours of the endocardium were automatically traced and manually corrected, if necessary.

Regional wall thickness data were categorized according to the AHA 16 segment model [22]. In both SHIP and KORA, LGE was assessed by two independent readers and in case of disagreement, examined by a third [23, 24]. Concentricity as a marker of LV remodeling was calculated as end-diastolic myocardial mass divided by end-diastolic volume.

### Covariate assessment

In both SHIP and KORA, clinical data were obtained at a central study center in standardized procedures conducted by trained staff [5, 20, 21, 25], see Supplementary Text S1 for details. In KORA, known prior CVD (myocardial infarction, stroke, revascularization) was an exclusion criterion for MRI participation [21]. In SHIP, some individuals had prior CVD, defined as self-reported myocardial infarction or stroke. We furthermore defined visible LGE on CMR as an indicator of prior CVD.

In individuals without prior CVD or visible LGE on CMR, CVD risk was estimated based on traditional cardiovascular risk factors, using the 10-year SCORE2 [26], 10-year Framingham Risk Score (FRS10 [27]), the 30-year Framingham Risk Score with competing risks (FRS30 [28]). Scores were calibrated to the actual sample, where applicable. We note that original outcomes for these scores vary, e.g. FRS10 includes non-fatal heart failure, whereas FRS30 and SCORE2 only include fatal heart failure.

### Incident outcome assessment

In SHIP, incident myocardial infarction, stroke and heart failure were ascertained at the follow-up examinations (SHIP-START-3/4, approximately 4-5 years after the MRI exam, and SHIP-TREND-1, approximately 7 years after the MRI exam, respectively). All-cause mortality was ascertained by linkage to physicians’ records, with a median follow-up time of 11 years. In KORA, incident myocardial infarction and mortality were ascertained at the follow-up examination in 2021/2022 (KORA-FFF4) or by physician death records, respectively. The main outcomes of interest were 1) the composite outcome of myocardial infarction, stroke, and heart failure morbidity with overall mortality, 2) the composite outcome of myocardial infarction, stroke, and heart failure morbidity with CVD mortality 3) overall mortality.

### Statistical methods

Missing values in clinical data were rare (<0.5%) and were imputed by random forest imputation based on a maximum of 10 iterations with 100 trees. Clinical and CMR data are presented as mean and standard deviation for continuous and counts and percentages for categorical variables. Differences between men and women were assessed with t-test and χ^2^ Test, respectively.

Phenotypes of LV remodeling patterns were identified by unsupervised k-means clustering with the appropriate number of clusters based on Calinski-Harabasz criterion. Individuals with prior CVD were kept in the sample for clustering, since we expected their remodeling characteristics to be valuable information for identifying individuals with similar remodeling, but without prior CVD. Resulting clusters were labeled with capital letters. As the main CMR parameter set for clustering (Set 1), we used the average wall thickness in each of the 16 AHA segments at both end-diastole and end-systole, resulting in 32 variables for clustering. For comparison, we used an additional set (Set 2) containing the 32 wall thickness variables plus ejection fraction, heart rate, and both end-diastolic and end-systolic volume and mass. Agreement between resulting clusters was calculated by transition probabilities. To account for inherent differences in MRI assessment between studies, CMR data were centered and scaled (minus mean and divided by standard deviation) stratified by sex and study before clustering.

Distribution of estimated CVD risk according to resulting remodeling phenotypes was visualized by boxplots. For comparison, distribution of risk according to quartiles of end-diastolic mass, average end-diastolic wall thickness and LV concentricity was visualized. Associations between remodeling phenotypes and CVD risk were evaluated by χ^2^ test on the distribution of individuals with LGE or prevalent CVD between clusters, and by linear regression models with outcome SCORE2, FRS10, and FRS30 for individuals without LGE/CVD. Confounder adjustment was as follows: 1) none, 2) end-diastolic mass, 3) average overall end-diastolic wall thickness, 4) concentricity, 5) concentricity and average overall wall thickness, 6) systolic and diastolic blood pressure. The first cluster level served as the reference category. Since CVD risk was estimated by traditional risk factors, further model adjustment for these risk factors was not sensible. R^2^ was used to denote the percentage of variance within the risk scores that could be explained by the model.

To identify relevant clinical variables associated with the distinct remodeling phenotypes, we used regularized multinomial logistic regression with LASSO penalty and symmetric parametrization for variable selection. The procedure was repeated on 1000 bootstrap samples of the data and the selection frequency of each variable served as a measure of relative importance.

Association of remodeling phenotypes with the incident outcomes of interest described above was assessed by logistic regression. Confounder adjustment was as follows: 1) none, 2) end-diastolic mass, 3) average overall end-diastolic wall thickness, 4) age, 5) SCORE2, FRS10, or FRS30. In the subset comprising only SHIP data (where mortality follow-up time was available), Kaplan-Meier Curves for all-cause mortality were plotted and compared by a weighted logrank permutation test [29]. Associations were evaluated by Cox proportional hazards regression, adjusted for age. See Supplementary Text S2 for additional details.

All analyses were stratified by sex. P-values <0.05 were considered to denote statistical significance, where applicable. We used R version 4.2.2 for all calculations.

## Results

### Study sample

The final sample comprised 991 men and 798 women, with the majority of participants originating from the SHIP-TREND-0 study (Figure S1, Table 1). Mean age in the sample was 52.7 years and prevalence of hypertension was 48.1% in men and 33.8% in women. Prior CVD, or LGE on CMR was present for 7.8% of men and 2.0% of women. For all others, estimated 10-year risk of CVD was 12.0% by FRS and 5.0% by SCORE2, with expected pronounced differences between men and women (Table 1).

**Table 1:** Participant characteristics.

|  | All | Men | Women | p-value |
| --- | --- | --- | --- | --- |
|  | N = 1789 | N = 991 (55.4%) | N = 798 (44.6%) |  |
| **General** |  |  |  |  |
| Age, years | 52.7 ± 12.7 | 52.9 ± 13.0 | 52.5 ± 12.3 | 0.492 |
| Study |  |  |  | 0.155 |
| SHIP-TREND-0 | 931 (52.0%) | 524 (52.9%) | 407 (51.0%) |  |
| SHIP-START-2 | 490 (27.4%) | 254 (25.6%) | 236 (29.6%) |  |
| KORA-FF4 | 368 (20.6%) | 213 (21.5%) | 155 (19.4%) |  |
| **Anthropometry** |  |  |  |  |
| Height, cm | 171.1 ± 9.4 | 176.9 ± 6.7 | 163.8 ± 6.7 | <0.001 |
| Weight, kg | 80.7 ± 15.2 | 87.7 ± 12.7 | 72.0 ± 13.5 | <0.001 |
| BMI, kg/m<sup>2</sup> | 27.5 ± 4.3 | 28.0 ± 3.7 | 26.8 ± 4.9 | <0.001 |
| Waist circumference, cm | 91.4 ± 13.2 | 96.8 ± 11.1 | 84.6 ± 12.4 | <0.001 |
| Hip circumference, cm | 102.2 ± 8.8 | 102.3 ± 7.3 | 102.0 ± 10.4 | 0.535 |
| **Lifestyle factors** |  |  |  |  |
| Smoking |  |  |  | <0.001 |
| Never | 683 (38.2%) | 316 (31.9%) | 367 (46.0%) |  |
| Former | 731 (40.9%) | 471 (47.5%) | 260 (32.6%) |  |
| Current | 375 (21.0%) | 204 (20.6%) | 171 (21.4%) |  |
| Physical Activity | 1249 (69.8%) | 669 (67.5%) | 580 (72.7%) | 0.021 |
| Alcohol consumption | 1581 (88.4%) | 903 (91.1%) | 678 (85.0%) | <0.001 |
| **Blood Pressure** |  |  |  |  |
| systolic, mmHg | 126.7 ± 17.7 | 132.6 ± 16.1 | 119.4 ± 16.9 | <0.001 |
| diastolic, mmHg | 78.1 ± 10.2 | 80.7 ± 10.1 | 74.9 ± 9.5 | <0.001 |
| Hypertension | 747 (41.8%) | 477 (48.1%) | 270 (33.8%) | <0.001 |
| Antihypertensive medication | 474 (26.5%) | 270 (27.2%) | 204 (25.6%) | 0.455 |
| **Glycemia** |  |  |  |  |
| HbA1c, % | 5.4 ± 0.7 | 5.4 ± 0.8 | 5.3 ± 0.6 | <0.001 |
| Fasting glucose, mmol/L | 5.7 ± 1.3 | 5.9 ± 1.4 | 5.4 ± 1.0 | <0.001 |
| Diabetes | 158 (8.8%) | 101 (10.2%) | 57 (7.1%) | 0.030 |
| **Lipid Profile** |  |  |  |  |
| Total Cholesterol, mmol/L | 5.5 ± 1.0 | 5.5 ± 1.0 | 5.6 ± 1.0 | 0.010 |
| Triglycerides, mmol/L | 1.6 ± 1.1 | 1.8 ± 1.3 | 1.3 ± 0.7 | <0.001 |
| HDL, mmol/L | 1.5 ± 0.4 | 1.3 ± 0.3 | 1.7 ± 0.4 | <0.001 |
| LDL, mmol/L | 3.5 ± 0.9 | 3.5 ± 0.9 | 3.4 ± 0.9 | 0.072 |
| Inflammation |  |  |  |  |
| Serum uric acid, $\mu\text{mol/L}$ | $296.2 \pm 81.0$ | $336.4 \pm 72.0$ | $246.4 \pm 61.6$ | $<0.001$ |
| Kidney |  |  |  |  |
| Creatinine, $\mu\text{mol/L}$ | $76.5 \pm 14.3$ | $84.2 \pm 11.9$ | $67.0 \pm 10.7$ | $<0.001$ |
| Cystatin C, $\text{mg/dL}$ | $0.7 \pm 0.1$ | $0.7 \pm 0.1$ | $0.7 \pm 0.1$ | $<0.001$ |
| eGFR, $\text{ml/min/1.73m}^2$ | $101.0 \pm 18.1$ | $102.3 \pm 18.0$ | $99.4 \pm 18.0$ | $<0.001$ |
| CVD |  |  |  |  |
| prior CVD, or LGE present on CMR | 93 (5.2%) | 77 (7.8%) | 16 (2.0%) | $<0.001$ |
| Risk scores (N=1696) |  |  |  |  |
| SCORE2, % | $5.0 \pm 4.3$ | $5.9 \pm 4.9$ | $4.0 \pm 3.3$ | $<0.001$ |
| FRS 10 years, % | $12.0 \pm 11.9$ | $15.9 \pm 13.6$ | $7.5 \pm 7.3$ | $<0.001$ |
| FRS 30 years, % | $15.4 \pm 12.7$ | $18.9 \pm 13.6$ | $11.3 \pm 10.0$ | $<0.001$ |
| Incident composite outcome | 205 (11.5%) | 141 (14.2%) | 64 (8.0%) | $<0.001$ |
| Incident CVD outcome | 142 (7.9%) | 99 (10.0%) | 43 (5.4%) | $<0.001$ |
| All-cause mortality | 85 (4.8%) | 62 (6.3%) | 23 (2.9%) | 0.001 |

Data are given as mean ± standard deviation or N(%). P-values from t-test or $\chi^2$ -Test, where applicable. LGE: Late Gadolinium Enhancement, FRS: Framingham Risk Score. Incident composite outcome denotes myocardial infarction, stroke, heart failure, and all-cause mortality. Incident CVD outcome denotes myocardial infarction, stroke, heart failure, and CVD mortality

On CMR, mean EF was 60.1%, mean EDV 146.2 ml, and average overall end-diastolic wall thickness 6.9 mm (Table S1).

### Phenotypes of LV remodeling patterns

Unsupervised clustering on wall thickness data for 16 AHA segments at end-diastole and end-systole resulted in k=4 clusters for both men and women. These were labeled cluster A to D and represented different phenotypes of LV remodeling. Clusters were characterized by gradually increasing wall thickness (Figure 1, respective upper panel) and myocardial mass, and gradually decreasing volumes (Table 2). Patterns for the distribution of EF, heart rate, cardiac output and stroke volume were less distinct (Table 2). In men, cardiac output and EF were comparable between clusters B and D, whereas in women, there was a gradual increase of these parameters from A-D. Stroke volume was highest in Cluster B in both men and women.

**Figure 1:**
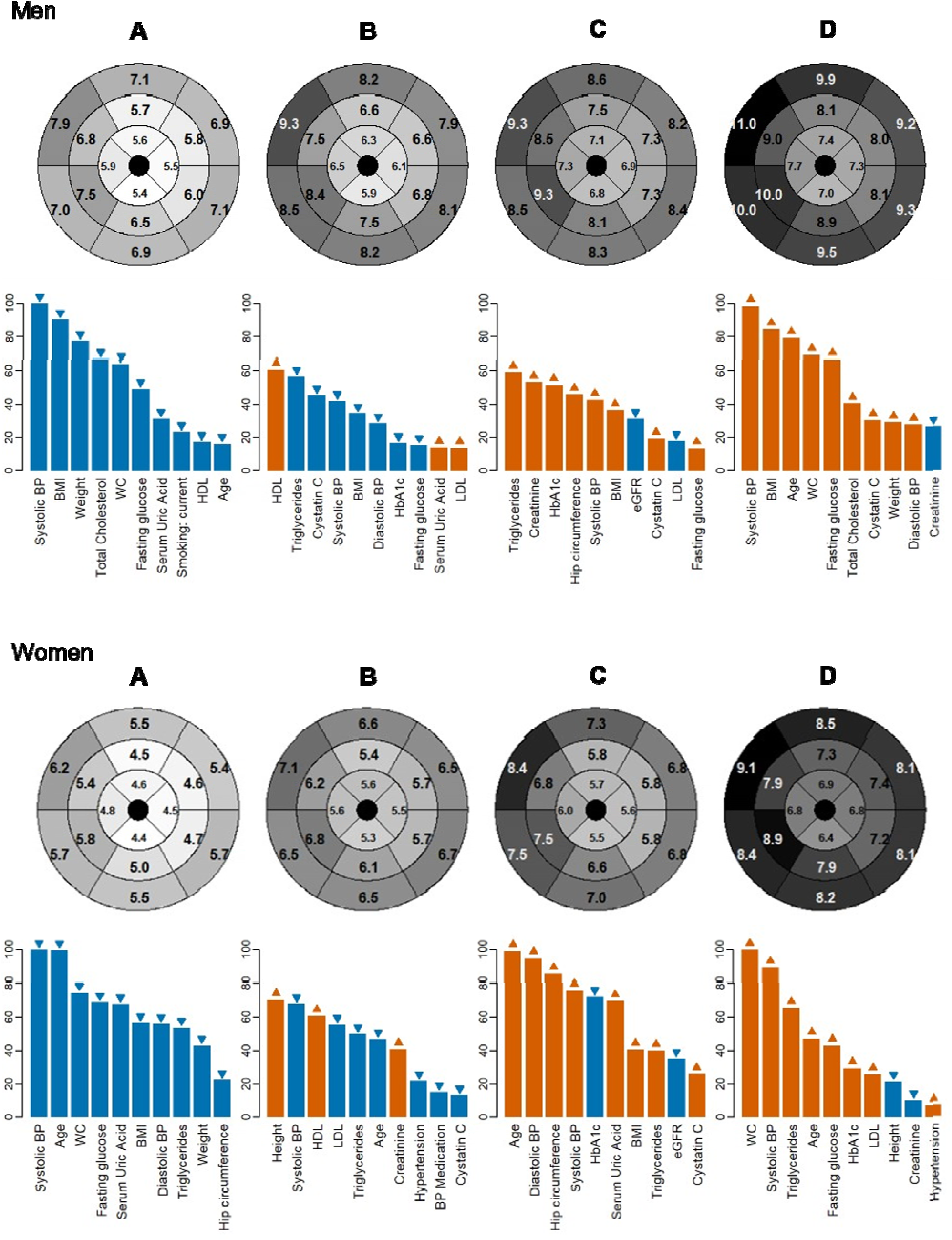
Wall thickness patterns in remodeling clusters, and their clinical determinants For men and women respectively: Upper panel: End-diastolic wall thickness per AHA segment in mm according to cluster A-D. Colors reflect relative wall thickness for each sex separately. Lower panel: Selection frequency (%, y-axis) of clinical determinants (x-axis) of each cluster A-D. Selection frequencies were derived from LASSO regularized multinomial regression with symmetric parametrization on 1000 bootstrap samples. Blue bars: Odds Ratio < 1 (increase of clinical variable decreases probability of remodeling pattern in cluster), Orange bars: Odds Ratio > 1 (increase of clinical variable increases probability of remodeling pattern in cluster). WC: waist circumference.

**Table 2:**
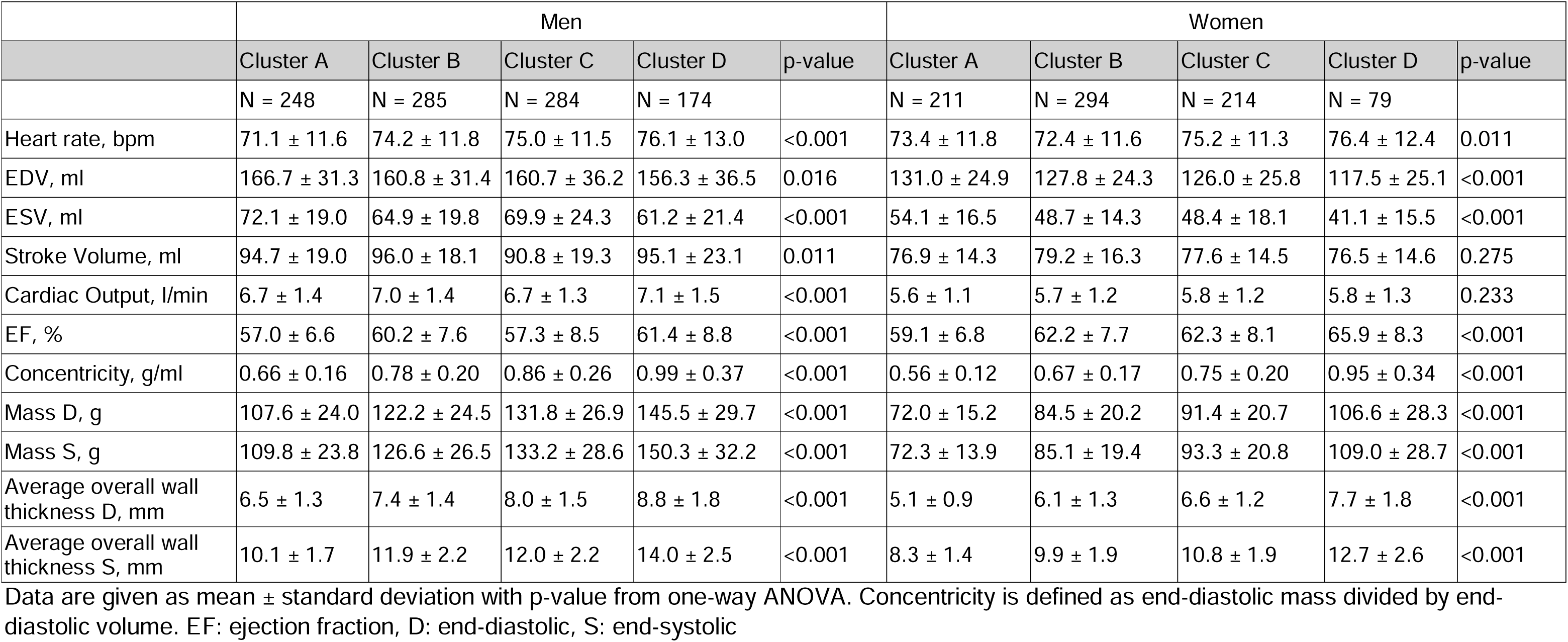
CMR parameters according to cluster of remodeling patter.

|  | Men |  |  |  |  | Women |  |  |  |  |
| --- | --- | --- | --- | --- | --- | --- | --- | --- | --- | --- |
|  | Cluster A | Cluster B | Cluster C | Cluster D | p-value | Cluster A | Cluster B | Cluster C | Cluster D | p-value |
|  | N = 248 | N = 285 | N = 284 | N = 174 |  | N = 211 | N = 294 | N = 214 | N = 79 |  |
| Heart rate, bpm | 71.1 ± 11.6 | 74.2 ± 11.8 | 75.0 ± 11.5 | 76.1 ± 13.0 | <0.001 | 73.4 ± 11.8 | 72.4 ± 11.6 | 75.2 ± 11.3 | 76.4 ± 12.4 | 0.011 |
| EDV, ml | 166.7 ± 31.3 | 160.8 ± 31.4 | 160.7 ± 36.2 | 156.3 ± 36.5 | 0.016 | 131.0 ± 24.9 | 127.8 ± 24.3 | 126.0 ± 25.8 | 117.5 ± 25.1 | <0.001 |
| ESV, ml | 72.1 ± 19.0 | 64.9 ± 19.8 | 69.9 ± 24.3 | 61.2 ± 21.4 | <0.001 | 54.1 ± 16.5 | 48.7 ± 14.3 | 48.4 ± 18.1 | 41.1 ± 15.5 | <0.001 |
| Stroke Volume, ml | 94.7 ± 19.0 | 96.0 ± 18.1 | 90.8 ± 19.3 | 95.1 ± 23.1 | 0.011 | 76.9 ± 14.3 | 79.2 ± 16.3 | 77.6 ± 14.5 | 76.5 ± 14.6 | 0.275 |
| Cardiac Output, l/min | 6.7 ± 1.4 | 7.0 ± 1.4 | 6.7 ± 1.3 | 7.1 ± 1.5 | <0.001 | 5.6 ± 1.1 | 5.7 ± 1.2 | 5.8 ± 1.2 | 5.8 ± 1.3 | 0.233 |
| EF, % | 57.0 ± 6.6 | 60.2 ± 7.6 | 57.3 ± 8.5 | 61.4 ± 8.8 | <0.001 | 59.1 ± 6.8 | 62.2 ± 7.7 | 62.3 ± 8.1 | 65.9 ± 8.3 | <0.001 |
| Concentricity, g/ml | 0.66 ± 0.16 | 0.78 ± 0.20 | 0.86 ± 0.26 | 0.99 ± 0.37 | <0.001 | 0.56 ± 0.12 | 0.67 ± 0.17 | 0.75 ± 0.20 | 0.95 ± 0.34 | <0.001 |
| Mass D, g | 107.6 ± 24.0 | 122.2 ± 24.5 | 131.8 ± 26.9 | 145.5 ± 29.7 | <0.001 | 72.0 ± 15.2 | 84.5 ± 20.2 | 91.4 ± 20.7 | 106.6 ± 28.3 | <0.001 |
| Mass S, g | 109.8 ± 23.8 | 126.6 ± 26.5 | 133.2 ± 28.6 | 150.3 ± 32.2 | <0.001 | 72.3 ± 13.9 | 85.1 ± 19.4 | 93.3 ± 20.8 | 109.0 ± 28.7 | <0.001 |
| Average overall wall thickness D, mm | 6.5 ± 1.3 | 7.4 ± 1.4 | 8.0 ± 1.5 | 8.8 ± 1.8 | <0.001 | 5.1 ± 0.9 | 6.1 ± 1.3 | 6.6 ± 1.2 | 7.7 ± 1.8 | <0.001 |
| Average overall wall thickness S, mm | 10.1 ± 1.7 | 11.9 ± 2.2 | 12.0 ± 2.2 | 14.0 ± 2.5 | <0.001 | 8.3 ± 1.4 | 9.9 ± 1.9 | 10.8 ± 1.9 | 12.7 ± 2.6 | <0.001 |

Data are given as mean ± standard deviation with p-value from one-way ANOVA. Concentricity is defined as end-diastolic mass divided by end-diastolic volume. EF: ejection fraction, D: end-diastolic, S: end-systolic

The distribution of prior CVD, or LGE on CMR differed significantly between clusters in men (4.8%, 4.9%, 9.5%, 13.8%, p-value χ^2^ = 0.001), but not in women (0.5%, 2.0%, 2.8%, 3.8%, p-value χ^2^ = 0.127). Estimated CVD risk was increasing in a gradual fashion along clusters (Figure 2), a pattern that was not present along quartiles of end-diastolic mass, average overall end-diastolic wall thickness, or concentricity (Figure S2). In particular, the highest quartile of these parameters did not identify a high-risk category (Figure S2). Clusters were significantly associated with estimated CVD risk in a gradual fashion (Figure 3 + Table S3): In men, the remodeling pattern in Cluster B compared to cluster A was associated with an increase of 3.9 percentage points (95%-CI [1.6, 6.1]) in FRS10, whereas the pattern in Cluster D was associated with an increase of 12.3 ([9.7, 15.0]) percentage points. In women, Cluster B was associated with an increase of 2.4 percentage points ([1.3, 3.6]), and Cluster D with an increase of 10.6 ([8.9, 12.3]) percentage points in FRS10. Overall, direction and significance of associations were consistent between the three outcomes SCORE2, FRS10, and FRS30. Explained variance was generally lowest for SCORE2. In men, remodeling phenotypes alone explained 9.8% of variance in FRS30 and 3.8% of variance in SCORE2, whereas in women, remodeling phenotypes explained 22.2% of variance in FRS30 and 13.9% of variance in SCORE2.

**Figure 2:**
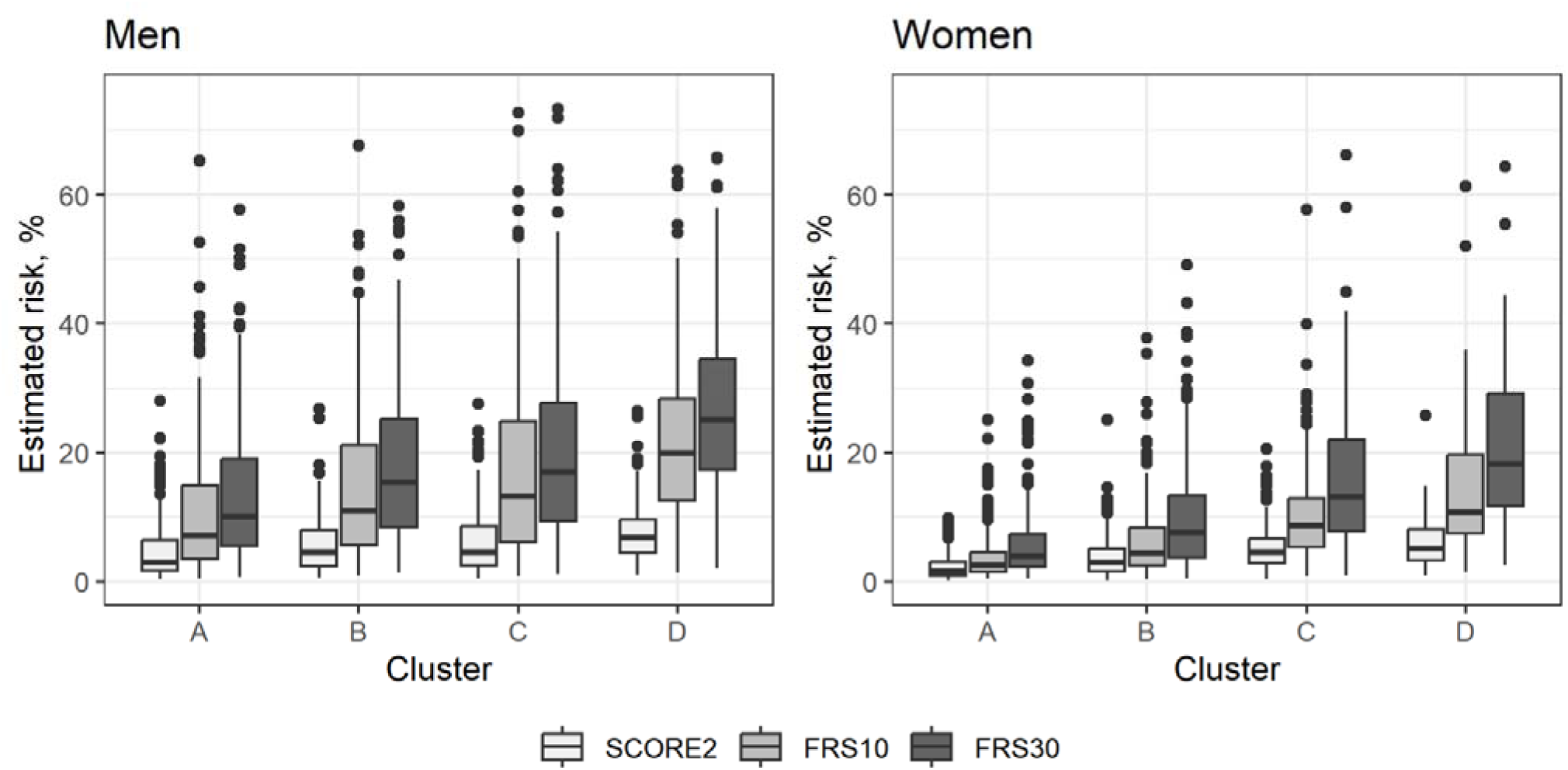
Distribution of estimated CVD risk according to clusters of remodeling patterns On the y-axis: Estimated CVD risk in %, on the x-axis: cluster of remodeling pattern

**Figure 3:**
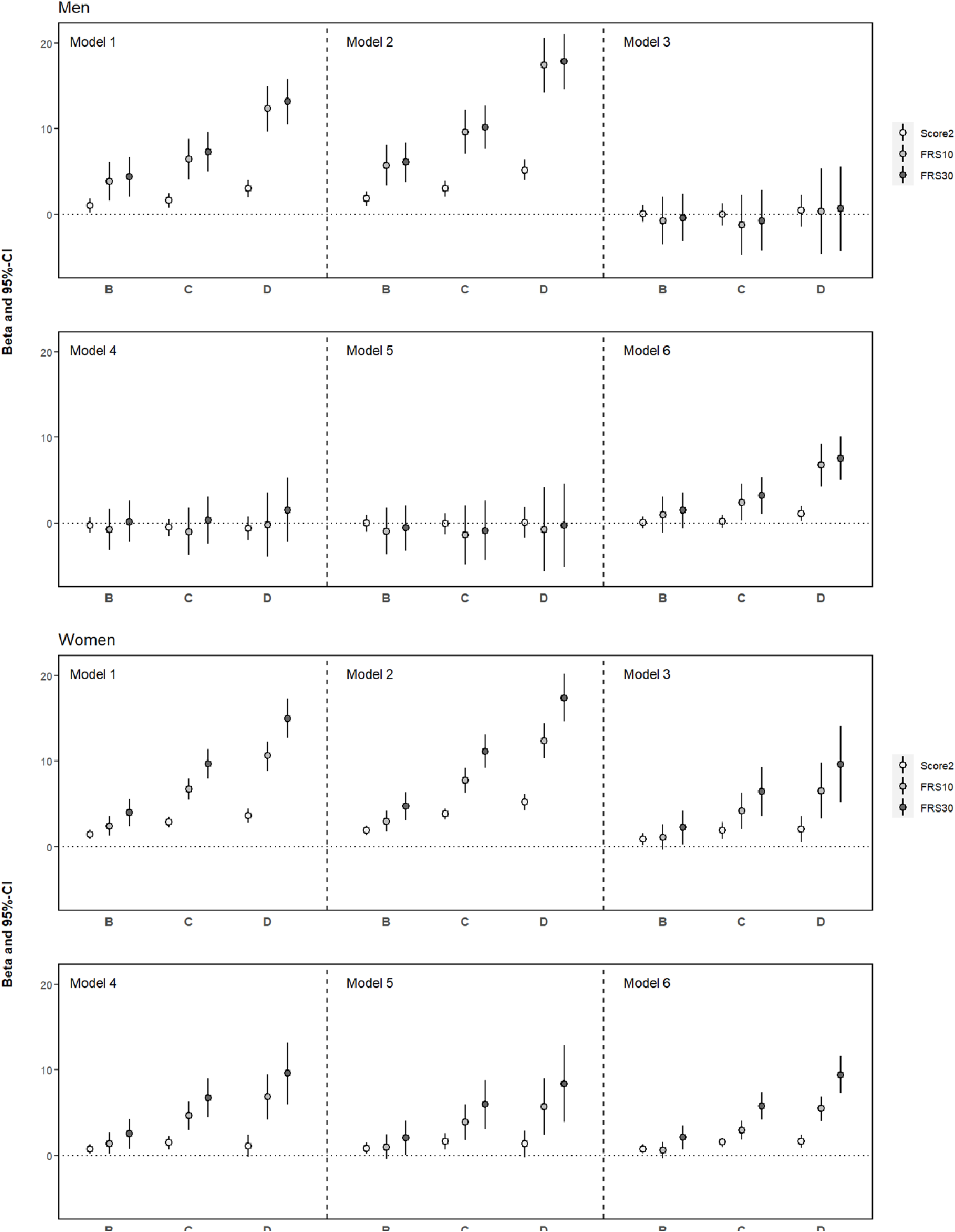
Associations of clusters of remodeling patterns with estimated CVD risk. On the y-axis: beta and 95%-CI from linear regression with exposure remodeling cluster (levels A, B, C, D) and outcome risk score (SCORE2: light grey, FRS10: medium grey, FRS30: dark grey). Cluster A served as the reference cluster. Adjustment was as follows: 1) none, 2) end-diastolic myocardial mass, 3) average overall end-diastolic wall thickness, 4) concentricity, 5) concentricity and average overall end-diastolic wall thickness, 6) systolic and diastolic blood pressure. Corresponding numerical data and R^2^ as measure of variance explained are presented in Table S3.

Associations of clusters of remodeling patterns were independent of myocardial mass (Figure 3 + Table S3, Model 2) and systolic and diastolic blood pressure (Figure 3 + Table S3, Model 6) for both men and women. However, only in women, associations of clusters with CVD risk were independent of average overall end-diastolic wall thickness (Figure 3 + Table S3, Model 3), concentricity (Figure 3 + Table S3, Model 4), and their combination (Figure 3 + Table S3, Model 5). Moreover in women, average overall wall thickness and concentricity added only marginally to the explained variance of the outcomes, e.g. 22.5% of variance explained in FRS10 compared to 21.2% from clusters alone (Table S3).

Between the two different CMR parameter sets used for clustering (Set 1: wall thickness at end-diastole and end-systole per AHA segment, Set 2: wall thickness per segment, EF, heart rate, volumes, mass at end-diastole and end-systole), transition probabilities ranged between 71.6% and 98.7% (Table S2). Most stable were the high-risk clusters C and D in women, which were mapped almost completely in the second parameter set. Associations of resulting clusters with CVD risk were similar when using Set 1 or Set 2 (Table S4); however effect sizes for women were consistently stronger when using Set 1.

### Clinical determinants of cluster of remodeling pattern

Clinical determinants of the respective clusters were identified from all variables in Table 1 (without study, CVD, and CVD risk scores) Automated variable selection based on regularized regression revealed that low-risk Cluster A was mainly characterized by an absence of traditional CVD risk factors (Figure 1, respective lower panel) in both men and women. Higher levels of systolic blood pressure, BMI and waist circumference, weight, or fasting glucose were associated with a lower probability of the wall thickness pattern in Cluster A. In women, younger age was more strongly associated with the low-risk remodeling pattern compared to men (selection frequency of 99.7% vs 16.7%).

Remodeling patterns of Cluster B in men were associated with a more favorable lipid profile and lower cystatin c, whereas remodeling in Cluster C was associated mainly with increased triglycerides, creatinine, and HbA1c. Generally, clinical determinants of intermediate risk Clusters B and C were less readily identified, as shown by lower selection frequencies (<65%). The high-risk remodeling pattern in Cluster D was mainly characterized by increased systolic blood pressure, BMI and WC, older age, and increased glucose.

In women, remodeling patterns in Cluster B were mainly associated with higher body height, lower blood pressure, and a more favorable lipid profile. Notably, women in Cluster B also had the lowest heart rate among clusters. Cluster C was mainly related to age and blood pressure. Interestingly, high diastolic blood pressure had a higher selection frequency and thus a higher relevance to the specific remodeling pattern in Cluster C. Furthermore, higher hip circumference and uric acid were positively associated with this remodeling pattern. High-risk Cluster D was characterized by higher waist circumference (selection frequency 100%), increased systolic blood pressure, and increased triglyceride levels (Figure 1, respective lower panel).

### Cluster of remodeling pattern and incident outcomes

Overall, there were 205 (11.5%) incident composite outcomes of myocardial infarction, stroke, heart failure and all-cause mortality, with expected differences between men and women (Table 1). Outcome distribution according to clusters of remodeling patterns was significantly different in men (p-value χ^2^ = 0.036), with the highest proportion of outcomes in Cluster D (Figure S3), but not in women (p-value χ^2^ = 0.179), with the highest proportion of outcomes in Cluster C (Figure S3).

In men, Cluster D was significantly associated with the incident composite outcome of myocardial infarction, stroke, heart failure, and mortality (OR 2.25, CI [1.29, 3.91], Table S5). In women, a similar pattern was seen for Cluster C (OR 2.09, CI [1.02, 4.31]). Associations for both men and women remained after adjustment for myocardial mass, but attenuated after adjustment for average overall wall thickness, age, or the FRS10 (Table S5). Similar patterns were observed for the composite CVD outcome of myocardial infarction, stroke, heart failure and CVD mortality (Table 1, Table S5).

In time-to-event analysis (Figure 4), survival distributions for men were significantly different in all clusters compared with Cluster A (all p<0.05), whereas for women, only Cluster C showed a significantly different survival distribution from Cluster A (p=0.011). In Cox regression adjusted for age, Cluster D in men had a higher hazard for all-cause mortality (HR 3.62, CI [1.33, 9.91]), whereas there was no significant association in women.

**Figure 4:**
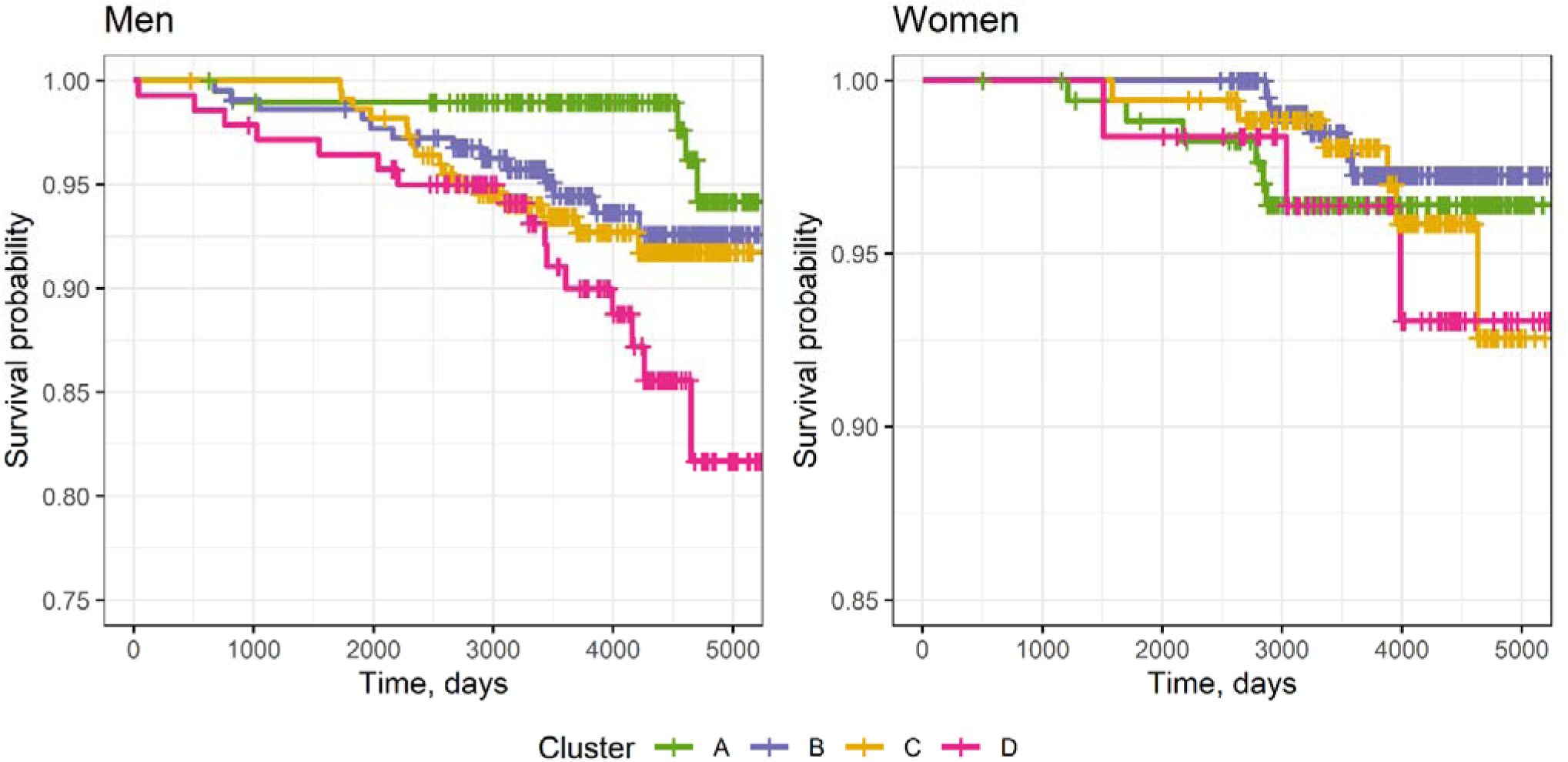
Survival probability according to cluster of remodeling pattern.

## Discussion

Using CMR data from three population-based studies, we found that regional patterns of left ventricular wall thickness cluster into distinct groups, and each group corresponds to a different level of estimated cardiovascular disease risk. Importantly, these risk groups could not be identified by analyzing just a single CMR parameter, such as myocardial mass, average wall thickness, or concentricity. Cardiac remodeling patterns were more strongly related to estimated CVD risk in women than men: they explained more variance of estimated CVD risk, and their association was more robust towards adjustment for single CMR parameters. While low-CVD risk patterns of remodeling were consistently associated with absence of traditional CVD risk factors, intermediate and high-risk remodeling patterns had different clinical determinants, including major differences between men and women.

The importance to characterize LV remodeling beyond mass and volumes has been recognized in previous population-based studies [1, 4, 5, 7, 8]. Recently, statistical shape features of LV remodeling, that have been developed in the MESA study, predicted incident CVD beyond traditional cardiovascular risk factors and single CMR parameters [13]. In this work, the authors emphasize the need to describe shape alterations within the regular AHA 16 segment classification to improve interpretability of the machine-learning derived signatures. However, to our knowledge, sex-specific morphometric cardiac atlases and subsequent statistical shape models have yet to be developed. In the current study, we found that remodeling patterns based on wall thickness data, according to the regular AHA 16 segment classification, identified individuals at high CVD risk, and showed even stronger associations when compared to patterns that were derived from wall thickness in addition to volumes, mass, EF, and heart rate, with a more pronounced effect in women.

Contrary to estimated CVD risk, we found no consistent gradual association of remodeling patterns with incident CVD morbidity and overall mortality; this association was notably absent in women. Besides statistical power considerations, this might be due to several reasons. First, we lacked the necessarily data granularity to differentiate causes of mortality. Neoplasms are the second most common cause of death in Germany, and we do not expect a strong relationship between the remodeling phenotypes and cancer mortality. Second, the greater proportion of variance explained in estimated CVD risk among women could also suggest that the remodeling patterns may be more closely related to underlying risk score components than to actual CVD events [16].

In this line, different remodeling patterns were associated with different clinical determinants. Consistent with the current knowledge about protective effects of female sex hormones, younger age was a major protective factor against unfavorable remodeling patterns in women, but less so in men. Estrogen not only has vasodilative effects and inhibits endothelial inflammation [30] but also slows the age-related decrease in viable cardiomyocytes [31]. Estrogen receptor beta signaling inhibits myocyte apoptosis and fibrosis [32], resulting in less hypertrophic remodeling in women. Moreover, estrogen fosters a more beneficial body composition characterized by predominant storage of adipose tissue in the gluteal and femoral regions. Conversely, the reduction of estrogen leads to a transition towards the buildup of visceral fat. In line with this, we found that in women, the high-risk pattern of LV remodeling was mainly characterized by increased waist circumference whereas an intermediate-risk pattern was mainly associated with increased hip circumference. In contrast, in men, BMI as a measure of general adiposity was the predominantly associated parameter for the high-risk cluster. Obesity has been previously reported to be a risk factor for LV remodeling on a population-based level [2, 24, 33] with different effects of subcutaneous and visceral adipose tissue; however, sex-specific analyses are scarce.

In women, remodeling patterns identified an intermediate risk Cluster B that was characterized by a low heart rate, a comparably favorable lipid profile, and higher body height. A protective effect of higher body height on coronary heart disease and heart failure has already been shown [34], which is probably due to decreased filling pressures and lower wall stress in taller individuals [35]. The beneficial effects of taller height could be partly modulated through genetically determined lipid profiles, as shared genetic variants have been identified resulting in a genetically more favorable lipid profile in taller people [36]. Our results add to these findings by introducing a specific LV remodeling pattern as an intermediate phenotype.

In men, Cluster C was characterized by increased wall thickness, normal EF, and reduced stroke volume. Conceptually, this corresponds to impaired longitudinal strain [37], whereas the profile of Cluster B with increased wall thickness, higher EF, and sustained stroke volume would point to regular longitudinal function [37]. Given the distribution of CVD risk estimated for these clusters, results are consistent with the association of longitudinal strain impairment with increased risk of adverse cardiac events and mortality [38].

Based on automated variable selection, lifestyle factors like smoking, physical activity, and alcohol consumption were considered less relevant for the remodeling patterns. Although the CVD population attributable fraction of smoking in Western Europe was recently reported to be more modest than previously thought [19], smoking had been found to be associated with less favorable LV remodeling patterns [39]. At the same time, analyses from the MESA study showed beneficial effects of physical activity on heart rate and LV morphology [40]. We hypothesize that in our analysis the impact of lifestyle factors was modulated through other biomarkers which were then more preferentially selected by the model, e.g. effects of smoking might have been captured by lipid profile or blood pressure [41, 42].

Hypertension has been established as the main risk factor of LV hypertrophy. In men, systolic blood pressure levels were the most relevant determinant of remodeling patterns: higher blood pressure was strongly associated with a high-CVD risk remodeling pattern, whereas lower blood pressure was strongly associated with a low-CVD risk remodeling pattern. Still, remodeling phenotypes were associated with estimated CVD risk even after adjustment for systolic and diastolic blood pressure, corroborating the complexity and multifactorial etiology of these states. Recent findings from the Framingham Study showed that effects of hypertension, diabetes, and obesity on echocardiographic measures of cardiac remodeling were partly synergistic and partly antagonistic [43]. Independent effects of glycemia on LV remodeling have been established [4, 5], however, insulin resistance might also act as a mediator in the association of adipose tissue and cardiac remodeling [44]. In our analysis, high fasting glucose levels were associated with high-risk clusters, and low levels with low-risk clusters in both men and women, but fasting glucose was less relevant for intermediate-risk clusters.

Interestingly, the intermediate risk Cluster C in women was more strongly associated with elevated diastolic than systolic blood pressure. A recent study from Japan including postmenopausal women reported that normal-to-high diastolic blood pressure was associated with diastolic, but not systolic dysfunction based on echocardiography measures [45], consistent with the increased prevalence of heart failure with preserved EF in that demographic group. However, an analysis of echocardiography data from three German population-based studies indicated that increased diastolic blood pressure was associated only with systolic dysfunction [46].

The current study has certain limitations. Given the non-granularity of morbidity and mortality data, further prospective studies with adequate follow-up time are needed to robustly investigate the predictive value of remodeling patterns on incident CVD. Moreover, our panel of clinical determinants lacked information on further potentially relevant parameters, such as coronary calcium, atrial fibrillation, angina pectoris, or genetic predisposition. Strengths of our analysis include the study population consisting of three population-based cohorts with cardiac magnetic resonance data, which enabled precise measurements of cardiac morphology and function, as well as the use of unbiased statistical methods for clustering and variable selection.

In conclusion, automating the acquisition of regional wall thickness data and integrating them with shape- or radiomics-based models could provide a promising tool for improved imaging-based CVD risk assessment for both men and women.

## Data Availability

The informed consent given by SHIP and KORA study participants does not cover data posting in public databases. However, data are available upon request through the official Transfer Portals by means of a project agreement, subject to approval by the Scientific Board. For SHIP, contact. For KORA, contact.

## Declarations

## CRediT author statement

Conceptualization: SR, TI, MRPM; Methodology: SR; Formal analysis: SR; Investigation: ChS, CS, RL, RB, MD, SBF, HV, AP, FB, CLS; Resources: ChS, CS, RL, RB, MD, SBF, CT, HV, AP, FB, CLS; Data Curation: SR, TI, MW, SS, MRPM; Writing - Original Draft: SR, TI, MW, MRPM; Writing - Review & Editing: SR, TI, MW, SS, ChS, CS, RL, RB, MD, SBF, CT, HV, AP, FB, CLS, MRPM; Visualization: SR, MW; Project Administration: SR, TI, SS, MRPM; Funding acquisition: SR, TI, MD, SBF, HV, AP, FB, CLS, MRPM

