## Supplementary Material for "Sex-Specific Patterns of Left Ventricular Remodeling Using Regional Wall Thickness Data and Their Associations with Cardiovascular Disease Risk"

Rospleszcz et al.

### Supplementary Texts

Supplementary Text S1: Clinical data assessment

**SHIP**

Generally, laboratory markers were obtained from venous blood samples in fasted condition that were drawn from each participant at the study center. All self-reports were obtained from standardized interviews conducted by trained staff.

*Anthropometry*

Weight was measured to the nearest 0.1 kg by calibrated digital scales while participants wore no shoes and light clothing. Waist circumference (WC) was measured to the nearest 0.1 cm with an inelastic tape between the lower rib margin and the iliac crest. Hip circumference was measured with an inelastic tape at the level of maximal gluteal protrusion.

*Lifestyle factors*

Smoking (never, former, or current) was defined according to self-report. Alcohol consumption (g/d) was calculated based on self-reported type and amount of alcoholic beverages consumed. Participants were categorized as physically active if they reported leisure time exercise for at least 1 h/week during summer or winter.

*Blood pressure*

After at least five minutes of resting, blood pressure was measured in seated position three times with an oscillometric digital device (OMRON HEM-705CP). The average of 2nd and 3rd measurement was taken as the final value. Hypertension was defined as blood pressure ≥140/90 or current antihypertensive treatment.

*Medication*

Intake of antihypertensive medication was assessed by self-report. Participants were asked to bring packages of their medications from the last 7 days before the interview. Antihypertensive medication was defined as medication including compounds classified by ATC-codes C02, C03, C07, C08, C09.

*Glycemia*

HbA1c was measured by high-performance liquid chromatography (Diamat, Bio-Rad Laboratories, Munich, Germany). Plasma fasting glucose was measured by a hexokinase method (Dimension Vista 1500, Siemens Healthcare Diagnostics, Eschborn, Germany). In SHIP-TREND-0, Diabetes was defined by physician-validated self-report or based on Oral Glucose Tolerance Test (2-hour glucose ≥200 mg/dL and/or fasting glucose >125 mg/dL). In SHIP-START-2, diabetes was defined by physician-validated self-report, HbA1c values ≥ 6.5% or fasting blood glucose ≥ 11.1 mol/l.

*Lipid Profile*

Lipids were measured photometrically (Dimension RxL or Dimension VISTA 1500, Siemens Healthcare Diagnostics, Eschborn, Germany).

*Other values*

Serum uric acid was measured photometrically (Dimension VISTA 1500, Siemens Healthcare Diagnostics, Eschborn, Germany). Creatinine was measured with a modified kinetic Jaffé method (Dimension RxL or Dimension Vista 1500, Siemens Healthcare Diagnostics, Eschborn, Germany).

Cystatin C was measured by nephelometric assays (BN ProSpec, Siemens Healthcare Diagnostics, Eschborn, Germany. eGFR was calculated in a sex-specific fashion according to CKD-EPI based on creatinine and cystatin c.

*CVD*

Prevalent CVD was defined as a self-report of a physician's diagnosis of myocardial infarction or stroke. Incident myocardial infarction, stroke and heart failure were ascertained at the follow-up examinations (SHIP-START-3/4, approximately 4-5 years after the MRI exam, and SHIP-TREND-1, approximately 7 years after the MRI exam, respectively). All-cause mortality was ascertained by linkage to physicians' records, with a median follow-up time of 11 years.

**KORA**

Generally, laboratory markers were obtained from venous blood samples in fasted condition that were drawn from each participant at the study center. Self-reports were obtained from standardized interviews conducted by trained staff.

*Anthropometry*

Weight was measured by calibrated steelyards or digital scales (SECA 635 or SECA 877 or SECA measuring station 285, Seca GmbH & Co, KG, Hamburg, Germany). Height was measured by a calibrated leveling bar (SECA 242, Seca GmbH & Co, KG, Hamburg, Germany). Waist circumference was measured with an inelastic tape at the level midway between the lower rib margin and the iliac crest. Hip circumference was measured with an inelastic tape at the level of maximal gluteal protrusion.

*Lifestyle factors*

Smoking (never, former or current) was defined according to self-report. Alcohol consumption (g/d) was calculated based on self-reported type and amount of alcoholic beverages consumed. Participants were categorized as physically active if they reported leisure time exercise for at least 1 h/week during summer or winter.

*Blood pressure*

After at least five minutes of resting, blood pressure was measured in seated position three times with an oscillometric digital device (OMRON HEM-705CP). The average of 2nd and 3rd measurement was taken as the final value. Hypertension was defined as blood pressure ≥140/90 or current antihypertensive treatment.

*Medication*

Intake of antihypertensive medication was assessed by self-report. Participants were asked to bring packages of their medications from the last 7 days before the interview. Antihypertensive medication was defined as medication including compounds classified as antihypertensively effective by the most recent guidelines (C02, C03, C07, C08, C09), while participants were aware of having hypertension.

*Glycemia*

HbA1c was measured with a cation-exchange high performance liquid chromatographic, photometric assay (VARIANT II TURBO Hemoglobin Testing System, Bio-Rad Laboratories Inc, Hercules, US). Fasting glucose was measured by a UV test using enzymatic reference method with hexokinase (Vista, Siemens or Cobas, Roche). Diabetes was defined by physician-validated self-report or based on Oral Glucose Tolerance Test (2-hour glucose ≥200 mg/dL and/or fasting glucose >125 mg/dL)

*Lipid Profile*

Lipid profile was measured by enzymatic, colorimetric Flex assays (CHOL, HDLC, LDLC, TRIG) (Vista, Siemens or Cobas, Roche) from serum.

*Other values*

Serum uric acid was measured by an enzymatic colorimetric UA Flex assay (Vista, Siemens or Cobas, Roche). Creatinine was measured by a kinetic colorimetric CREJ assay based on Jaffé method. Cystatin C was measured by particle-enhanced immunonephelometry. eGFR was calculated in a sex-specific fashion according to CKD-EPI based on creatinine and cystatin c.

*CVD*

Per inclusion criteria, participants in KORA-MRI had no previous CVD. Incident myocardial infarction and mortality were ascertained at the follow-up examination in 2021/2022 (KORA-FFF4) or by physician death records, respectively. The main outcomes of interest were 1) the composite outcome of myocardial infarction, stroke, and heart failure morbidity with all-cause mortality, 2) all-cause mortality.

Supplementary Text S2: Analysis of incident outcomes.

For the main analysis, we combined incident morbidity and all-cause mortality to a composite outcome, and analyzed the composite outcome by logistic regression. The reason for combining morbidity and all-cause mortality was to maximize statistical power. Causes of death were available only for a subset of cases in SHIP, and not available at all for cases in KORA, which is why we could not focus on CVD mortality. By investigating all-cause mortality, our estimates will likely be biased towards the Null, since our hypothesis does not postulate associations between the remodeling phenotypes and e.g. cancer mortality. The reason for using bar plots and logistic regression, neglecting the time component, instead of Kaplan-Meier curves and Cox regression for the composite outcome was that time of event was not available for incident morbidity in neither SHIP nor KORA, and time of death was only available in SHIP. Thus, we used Kaplan Meier Curves only for the secondary mortality outcome in SHIP. Moreover, we saw (main Figure 4) that the Kaplan Meier Curves according to remodeling cluster partly crossed, violating the assumptions of the regular logrank test, so we used a weighted logrank permutation test which accounts for this situation.

### Supplementary Tables

Supplementary Table S1: CMR parameters of participants

| | All | Men | Women | p-value |
|---|---|---|---|---|
| | N = 1789 | N = 991 (55.4%) | N = 798 (44.6%) | |
| Heart rate, bpm | 73.9 ± 11.9 | 74.0 ± 12.0 | 73.8 ± 11.7 | 0.739 |
| Cardiac Output, l/min | 6.3 ± 1.4 | 6.9 ± 1.4 | 5.7 ± 1.2 | <0.001 |
| EDV, ml | 146.2 ± 34.8 | 161.5 ± 33.9 | 127.1 ± 25.2 | <0.001 |
| ESV, ml | 59.4 ± 21.5 | 67.5 ± 21.6 | 49.3 ± 16.5 | <0.001 |
| EF, % | 60.1 ± 8.1 | 58.8 ± 8.1 | 61.8 ± 7.9 | <0.001 |
| Myocardial ass D, g | 107.5 ± 33.0 | 125.4 ± 29.0 | 85.2 ± 22.5 | <0.001 |
| Myocardial mass S, g | 109.6 ± 34.4 | 128.5 ± 30.6 | 86.3 ± 22.4 | <0.001 |
| Stroke Volume, ml | 86.8 ± 19.5 | 94.0 ± 19.7 | 77.9 ± 15.2 | <0.001 |
| Wall thickness | | | | |
| Average overall D, mm | 6.9 ± 1.7 | 7.6 ± 1.7 | 6.1 ± 1.5 | <0.001 |
| Average overall S, mm | 11.0 ± 2.5 | 11.8 ± 2.5 | 10.0 ± 2.2 | <0.001 |
| Average basal D, mm | 7.6 ± 1.8 | 8.4 ± 1.7 | 6.7 ± 1.5 | <0.001 |
| Average basal S, mm | 11.2 ± 2.2 | 12.0 ± 2.1 | 10.2 ± 2.0 | <0.001 |
| Average mid D, mm | 6.8 ± 1.9 | 7.5 ± 1.9 | 6.0 ± 1.6 | <0.001 |
| Average mid S, mm | 11.5 ± 3.2 | 12.3 ± 3.2 | 10.4 ± 2.8 | <0.001 |
| Average apical D, mm | 6.0 ± 1.6 | 6.5 ± 1.6 | 5.4 ± 1.5 | <0.001 |
| Average apical S, mm | 10.0 ± 2.8 | 10.7 ± 2.8 | 9.1 ± 2.5 | <0.001 |
| Segment 1 D, mm | 7.6 ± 2.1 | 8.3 ± 2.1 | 6.7 ± 1.7 | <0.001 |
| Segment 1 S, mm | 11.6 ± 2.7 | 12.4 ± 2.6 | 10.5 ± 2.5 | <0.001 |
| Segment 2 D, mm | 8.4 ± 1.8 | 9.2 ± 1.7 | 7.4 ± 1.5 | <0.001 |
| Segment 2 S, mm | 11.1 ± 2.6 | 11.9 ± 2.5 | 10.1 ± 2.3 | <0.001 |
| Segment 3 D, mm | 7.7 ± 1.7 | 8.4 ± 1.6 | 6.7 ± 1.3 | <0.001 |
| Segment 3 S, mm | 10.1 ± 2.2 | 10.9 ± 2.2 | 9.2 ± 1.8 | <0.001 |
| Segment 4 D, mm | 7.4 ± 2.1 | 8.1 ± 2.0 | 6.5 ± 1.8 | <0.001 |
| Segment 4 S, mm | 11.1 ± 2.4 | 11.9 ± 2.3 | 10.1 ± 2.0 | <0.001 |
| Segment 5 D, mm | 7.5 ± 2.2 | 8.2 ± 2.2 | 6.6 ± 2.1 | <0.001 |
| Segment 5 S, mm | 11.8 ± 2.8 | 12.6 ± 2.8 | 10.7 ± 2.5 | <0.001 |
| Segment 6 D, mm | 7.3 ± 2.1 | 8.0 ± 2.1 | 6.4 ± 1.9 | <0.001 |
| Segment 6 S, mm | 11.5 ± 2.9 | 12.4 ± 2.8 | 10.5 ± 2.6 | <0.001 |
| Segment 7 D, mm | 6.2 ± 2.0 | 6.9 ± 2.1 | 5.5 ± 1.7 | <0.001 |
| Segment 7 S, mm | 11.1 ± 3.8 | 11.9 ± 3.8 | 10.0 ± 3.4 | <0.001 |
| Segment 8 D, mm | 7.2 ± 1.8 | 7.9 ± 1.9 | 6.3 ± 1.4 | <0.001 |
| Segment 8 S, mm | 11.7 ± 2.7 | 12.5 ± 2.6 | 10.7 ± 2.4 | <0.001 |
| Segment 9 D, mm | 7.9 ± 2.0 | 8.7 ± 1.9 | 6.9 ± 1.6 | <0.001 |
| Segment 9 S, mm | 12.1 ± 2.5 | 13.0 ± 2.4 | 11.0 ± 2.2 | <0.001 |
| Segment 10 D, mm | 7.0 ± 2.1 | 7.7 ± 2.1 | 6.1 ± 1.8 | <0.001 |
| Segment 10 S, mm | 11.4 ± 3.2 | 12.2 ± 3.2 | 10.4 ± 3.0 | <0.001 |
| Segment 11 D, mm | 6.4 ± 2.0 | 7.0 ± 2.0 | 5.6 ± 1.8 | <0.001 |
| Segment 11 S, mm | 11.3 ± 3.7 | 12.1 ± 3.8 | 10.3 ± 3.4 | <0.001 |
| Segment 12 D, mm | 6.3 ± 2.0 | 6.9 ± 2.0 | 5.6 ± 1.8 | <0.001 |
| Segment 12 S, mm | 11.3 ± 3.9 | 12.1 ± 4.0 | 10.3 ± 3.5 | <0.001 |
| Segment 13 D, mm | 6.1 ± 1.8 | 6.5 ± 1.9 | 5.5 ± 1.6 | <0.001 |
| Segment 13 S, mm | 10.1 ± 3.3 | 10.9 ± 3.4 | 9.1 ± 2.9 | <0.001 |
| Segment 14 D, mm | 6.3 ± 1.5 | 6.8 ± 1.4 | 5.6 ± 1.3 | <0.001 |
| Segment 14 S, mm | 10.3 ± 2.4 | 11.0 ± 2.4 | 9.4 ± 2.1 | <0.001 |

| | | | | |
|---|---|---|---|---|
| Segment 15 D, mm | 5.8 ± 1.6 | 6.2 ± 1.5 | 5.2 ± 1.5 | <0.001 |
| Segment 15 S, mm | 9.5 ± 2.6 | 10.2 ± 2.6 | 8.7 ± 2.4 | <0.001 |
| Segment 16 D, mm | 5.9 ± 1.9 | 6.4 ± 2.0 | 5.4 ± 1.8 | <0.001 |
| Segment 16 S, mm | 10.1 ± 3.4 | 10.9 ± 3.5 | 9.1 ± 3.1 | <0.001 |

Data are given as mean ± standard deviation with P-values from t-test. EDV: End diastolic volume, ESV: End systolic volume, EF: Ejection Fraction, D: end-diastolic, S: end-systolic.

Supplementary Table S2: Transition probabilities from clusters based on Set 1 to clusters based on Set 2

| Men | | | | | |
|---|---|---|---|---|---|
| | | Clustering based on Set 2 | | | |
| | | A | B | C | D |
| Clustering based on Set 1 | A | 84.3% | 15.7% | 0.0% | 0.0% |
| | B | 4.6% | 71.6% | 22.5% | 1.4% |
| | C | 0.4% | 12.0% | 83.8% | 3.9% |
| | D | 0.0% | 0.0% | 8.0% | 92.0% |
| Women | | | | | |
| | | Clustering based on Set 1 | | | |
| | | A | B | C | D |
| Clustering based on Set 1 | A | 72.5% | 27.5% | 0.0% | 0.0% |
| | B | 1.0% | 74.8% | 24.1% | 0.0% |
| | C | 0.0% | 1.9% | 95.3% | 2.8% |
| | D | 0.0% | 0.0% | 1.3% | 98.7% |

Set 1 contained wall thickness data for 16 AHA segments at end-diastole and end-systole. Set 2 contained the same wall thickness data, plus ejection fraction, heart rate, end-diastolic and end-systolic volume, and myocardial mass. Percentages denote the proportion of individuals in each level from clusters based on Set 1 that transition to the respective level from clusters based on Set 2. Therefore, percentages add up to 100% per row.

Supplementary Table S3: Associations of clusters of remodeling patterns with estimated CVD risk.

| | | Men | | | | | | | | | Women | | | | | | | | |
|---|---|---|---|---|---|---|---|---|---|---|---|---|---|---|---|---|---|---|---|
| | | SCORE2 | | | FRS10 | | | FRS30 | | | SCORE2 | | | FRS10 | | | FRS30 | | |
| | | beta | 95% CI | p-value | beta | 95% CI | p-value | beta | 95% CI | p-value | beta | 95% CI | p-value | beta | 95% CI | p-value | beta | 95 %CI | p-value |
| Model 1 | B | 1.07 | [0.2, 1.9] | 0.012 | 3.85 | [1.6, 6.1] | 0.001 | 4.40 | [2.1, 6.7] | <0.001 | 1.43 | [0.9, 2.0] | <0.001 | 2.43 | [1.3, 3.6] | <0.001 | 3.99 | [2.4, 5.6] | <0.001 |
| | C | 1.64 | [0.8, 2.5] | <0.001 | 6.45 | [4.1, 8.8] | <0.001 | 7.26 | [5.0, 9.6] | <0.001 | 2.88 | [2.3, 3.5] | <0.001 | 6.71 | [5.5, 8.0] | <0.001 | 9.70 | [8.0, 11.4] | <0.001 |
| | D | 3.02 | [2.0, 4.0] | <0.001 | 12.34 | [9.7, 15.0] | <0.001 | 13.19 | [10.5, 15.8] | <0.001 | 3.65 | [2.8, 4.5] | <0.001 | 10.63 | [8.9, 12.3] | <0.001 | 14.99 | [12.7, 17.3] | <0.001 |
| | $R^2$ | | | 0.038 | | | 0.085 | | | 0.098 | | | 0.139 | | | 0.212 | | | 0.222 |
| Model 2 | B | 1.87 | [1.0, 2.7] | <0.001 | 5.71 | [3.4, 8.1] | <0.001 | 6.09 | [3.8, 8.4] | <0.001 | 1.93 | [1.4, 2.5] | <0.001 | 2.98 | [1.8, 4.2] | <0.001 | 4.75 | [3.1, 6.4] | <0.001 |
| | C | 3.01 | [2.1, 3.9] | <0.001 | 9.65 | [7.1, 12.2] | <0.001 | 10.18 | [7.6, 12.7] | <0.001 | 3.83 | [3.2, 4.5] | <0.001 | 7.76 | [6.3, 9.2] | <0.001 | 11.15 | [9.2, 13.1] | <0.001 |
| | D | 5.19 | [4.0, 6.4] | <0.001 | 17.42 | [14.2, 20.6] | <0.001 | 17.82 | [14.6, 21.0] | <0.001 | 5.22 | [4.3, 6.2] | <0.001 | 12.36 | [10.3, 14.4] | <0.001 | 17.39 | [14.6, 20.2] | <0.001 |
| | $R^2$ | | | 0.077 | | | 0.112 | | | 0.120 | | | 0.172 | | | 0.219 | | | 0.230 |
| Model 3 | B | 0.1 | [-0.9, 1.1] | 0.846 | -0.71 | [-3.5, 2.1] | 0.614 | -0.38 | [-3.1, 2.4] | 0.784 | 0.92 | [0.2, 1.6] | 0.009 | 1.11 | [-0.3, 2.6] | 0.134 | 2.27 | [0.3, 4.2] | 0.025 |
| | C | 0.01 | [-1.3, 1.3] | 0.982 | -1.20 | [-4.7, 2.3] | 0.507 | -0.74 | [-4.2, 2.8] | 0.679 | 1.93 | [0.9, 2.9] | <0.001 | 4.22 | [2.1, 6.3] | <0.001 | 6.45 | [3.6, 9.3] | <0.001 |
| | D | 0.48 | [-1.4, 2.3] | 0.609 | 0.39 | [-4.6, 5.4] | 0.878 | 0.68 | [-4.3, 5.6] | 0.787 | 2.07 | [0.5, 3.6] | 0.009 | 6.51 | [3.3, 9.8] | <0.001 | 9.62 | [5.2, 14.1] | <0.001 |
| | $R^2$ | | | 0.047 | | | 0.114 | | | 0.130 | | | 0.144 | | | 0.219 | | | 0.229 |
| Model 4 | B | -0.24 | [-1.1, 0.7] | 0.602 | -0.70 | [-3.1, 1.7] | 0.567 | 0.16 | [-2.2, 2.6] | 0.894 | 0.75 | [0.2, 1.3] | 0.013 | 1.42 | [0.2, 2.7] | 0.028 | 2.54 | [0.8, 4.3] | 0.004 |
| | C | -0.49 | [-1.5, 0.5] | 0.342 | -0.98 | [-3.7, 1.8] | 0.483 | 0.36 | [-2.4, 3.1] | 0.798 | 1.52 | [0.7, 2.3] | <0.001 | 4.66 | [3.0, 6.3] | <0.001 | 6.76 | [4.5, 9.0] | <0.001 |
| | D | -0.58 | [-2.0, 0.8] | 0.418 | -0.18 | [-3.9, 3.6] | 0.926 | 1.56 | [-2.2, 5.3] | 0.415 | 1.14 | [-0.1, 2.4] | 0.068 | 6.86 | [4.2, 9.5] | <0.001 | 9.59 | [6.0, 13.2] | <0.001 |
| | $R^2$ | | | 0.085 | | | 0.158 | | | 0.161 | | | 0.167 | | | 0.224 | | | 0.236 |
| Model 5 | B | 0.04 | [-1.0, 1.0] | 0.943 | -0.91 | [-3.6, 1.8] | 0.510 | -0.55 | [-3.2, 2.1] | 0.688 | 0.83 | [0.2, 1.5] | 0.017 | 1.01 | [-0.4, 2.5] | 0.173 | 2.11 | [0.1, 4.1] | 0.036 |
| | C | -0.03 | [-1.3, 1.2] | 0.964 | -1.33 | [-4.8, 2.1] | 0.449 | -0.85 | [-4.3, 2.6] | 0.626 | 1.66 | [0.7, 2.6] | 0.001 | 3.92 | [1.8, 6.0] | <0.001 | 5.98 | [3.1, 8.8] | <0.001 |
| | D | 0.12 | [-1.7, 1.9] | 0.898 | -0.70 | [-5.6, 4.2] | 0.779 | -0.25 | [-5.1, 4.6] | 0.919 | 1.36 | [-0.2, 2.9] | 0.085 | 5.70 | [2.4, 9.0] | 0.001 | 8.38 | [3.9, 12.9] | <0.001 |
| | $R^2$ | | | 0.086 | | | 0.157 | | | 0.161 | | | 0.166 | | | 0.225 | | | 0.236 |
| Model 6 | B | 0.1 | [-0.6, 0.8] | 0.790 | 1.01 | [-1.1, 3.1] | 0.338 | 1.53 | [-0.6, 3.6] | 0.152 | 0.81 | [0.3, 1.3] | 0.001 | 0.63 | [-0.3, 1.6] | 0.191 | 2.12 | [0.7, 3.5] | 0.003 |
| | C | 0.26 | [-0.5, 1.0] | 0.509 | 2.43 | [0.3, 4.6] | 0.026 | 3.22 | [1.1, 5.4] | 0.004 | 1.57 | [1.0, 2.1] | <0.001 | 2.98 | [1.9, 4.1] | <0.001 | 5.80 | [4.2, 7.4] | <0.001 |
| | D | 1.13 | [0.2, 2.0] | 0.014 | 6.78 | [4.3, 9.3] | <0.001 | 7.56 | [5.0, 10.1] | <0.001 | 1.66 | [0.9, 2.4] | <0.001 | 5.50 | [4.0, 6.9] | <0.001 | 9.42 | [7.3, 11.6] | <0.001 |
| | $R^2$ | | | 0.273 | | | 0.272 | | | 0.262 | | | 0.393 | | | 0.497 | | | 0.413 |

Beta denotes coefficient from linear regression with exposure remodeling cluster (levels B, C, D, with A as the reference level) and outcome risk score (SCORE2, FRS10, FRS30). Adjustment was as follows: 1) none, 2) end-diastolic mass, 3) average overall end-diastolic wall thickness, 4) concentricity, 5) concentricity and average overall end-diastolic wall thickness, 6) systolic and diastolic blood pressure. $R^2$ denotes percentage of variance in risk scores explained.

Supplementary Table S4: Associations of clusters of remodeling patterns with estimated CVD risk when variable Set 2 is used for clustering.

| | | Men | | | | | | | | | Women | | | | | | | | |
|---|---|---|---|---|---|---|---|---|---|---|---|---|---|---|---|---|---|---|---|
| | | SCORE2 | | | FRS10 | | | FRS30 | | | SCORE2 | | | FRS10 | | | FRS30 | | |
| | | beta | 95% CI | p-value | beta | 95% CI | p-value | beta | 95% CI | p-value | beta | 95% CI | p-value | beta | 95% CI | p-value | beta | 95 %CI | p-value |
| Model 1 | B | 0.47 | [-0.4, 1.3] | 0.286 | 2.24 | [-0.1, 4.6] | 0.063 | 2.73 | [0.4, 5.1] | 0.023 | 1.57 | [1.0, 2.2] | 0.000 | 2.48 | [1.2, 3.8] | 0.000 | 4.11 | [2.3, 5.9] | 0.000 |
| | C | 0.94 | [0.1, 1.8] | 0.030 | 5.49 | [3.2, 7.8] | 0.000 | 6.55 | [4.2, 8.8] | 0.000 | 2.66 | [2.0, 3.3] | 0.000 | 5.86 | [4.6, 7.2] | 0.000 | 8.58 | [6.8, 10.3] | 0.000 |
| | D | 2.85 | [1.9, 3.8] | 0.000 | 12.25 | [9.5, 15.0] | 0.000 | 13.24 | [10.5, 15.9] | 0.000 | 3.76 | [2.9, 4.6] | 0.000 | 11.02 | [9.3, 12.8] | 0.000 | 15.69 | [13.3, 18.1] | 0.000 |
| | $R^2$ | | | 0.034 | | | 0.086 | | | 0.101 | | | 0.118 | | | 0.192 | | | 0.202 |
| Model 2 | B | 1.45 | [0.5, 2.4] | 0.002 | 4.75 | [2.3, 7.2] | 0.000 | 5.12 | [2.6, 7.6] | 0.000 | 1.94 | [1.3, 2.6] | 0.000 | 2.96 | [1.7, 4.3] | 0.000 | 4.76 | [3.0, 6.5] | 0.000 |
| | C | 2.67 | [1.7, 3.7] | 0.000 | 9.95 | [7.2, 12.7] | 0.000 | 10.78 | [8.0, 13.5] | 0.000 | 3.77 | [3.1, 4.5] | 0.000 | 7.29 | [5.8, 8.8] | 0.000 | 10.52 | [8.4, 12.6] | 0.000 |
| | D | 5.03 | [3.8, 6.3] | 0.000 | 17.84 | [14.5, 21.2] | 0.000 | 18.56 | [15.2, 21.9] | 0.000 | 5.54 | [4.5, 6.6] | 0.000 | 13.31 | [11.1, 15.5] | 0.000 | 18.80 | [15.8, 21.8] | 0.000 |
| | $R^2$ | | | 0.068 | | | 0.114 | | | 0.126 | | | 0.152 | | | 0.203 | | | 0.213 |
| Model 3 | B | -0.78 | [-1.8, 0.2] | 0.128 | -2.14 | [-4.9, 0.6] | 0.124 | -1.60 | [-4.3, 1.1] | 0.244 | 0.85 | [0.1, 1.5] | 0.017 | 0.98 | [-0.5, 2.5] | 0.191 | 2.08 | [0.1, 4.1] | 0.043 |
| | C | -1.59 | [-2.9, -0.2] | 0.0213 | -3.38 | [-7.0, 0.3] | 0.071 | -2.24 | [-5.9, 1.4] | 0.228 | 1.03 | [0.0, 2.0] | 0.043 | 2.47 | [0.4, 4.6] | 0.022 | 3.99 | [1.1, 6.9] | 0.007 |
| | D | -0.86 | [-2.7, 1.0] | 0.361 | -0.76 | [-5.8, 4.2] | 0.765 | 0.35 | [-4.6, 5.3] | 0.891 | 1.04 | [-0.5, 2.6] | 0.192 | 5.34 | [2.0, 8.7] | 0.002 | 8.02 | [3.5, 12.5] | 0.001 |
| | $R^2$ | | | 0.056 | | | 0.120 | | | 0.134 | | | 0.135 | | | 0.207 | | | 0.216 |
| Model 4 | B | -0.62 | [-1.5, 0.3] | 0.17 | -1.26 | [-3.7, 1.1] | 0.300 | -0.47 | [-2.9, 1.9] | 0.698 | 0.87 | [0.2, 1.5] | 0.007 | 1.38 | [0.0, 2.7] | 0.045 | 2.53 | [0.7, 4.4] | 0.007 |
| | C | -1.21 | [-2.2, -0.2] | 0.0175 | -1.40 | [-4.1, 1.3] | 0.307 | 0.24 | [-2.4, 2.9] | 0.858 | 1.15 | [0.4, 1.9] | 0.003 | 3.48 | [1.9, 5.1] | 0.000 | 5.19 | [3.0, 7.4] | 0.000 |
| | D | -1.03 | [-2.4, 0.4] | 0.148 | -0.24 | [-4.0, 3.5] | 0.901 | 1.82 | [-2.0, 5.6] | 0.344 | 0.87 | [-0.3, 2.1] | 0.156 | 6.45 | [3.9, 9.0] | 0.000 | 9.17 | [5.7, 12.7] | 0.000 |
| | $R^2$ | | | 0.090 | | | 0.160 | | | 0.162 | | | 0.162 | | | 0.214 | | | 0.226 |
| Model 5 | B | -0.62 | [-1.6, 0.4] | 0.222 | -1.64 | [-4.3, 1.0] | 0.226 | -1.18 | [-3.8, 1.5] | 0.384 | 0.82 | [0.1, 1.5] | 0.019 | 0.95 | [-0.5, 2.4] | 0.204 | 2.03 | [0.0, 4.0] | 0.046 |
| | C | -1.20 | [-2.5, 0.1] | 0.0795 | -2.18 | [-5.8, 1.4] | 0.237 | -1.22 | [-4.8, 2.4] | 0.508 | 1.05 | [0.1, 2.0] | 0.037 | 2.49 | [0.4, 4.6] | 0.020 | 4.02 | [1.2, 6.9] | 0.006 |
| | D | -1.02 | [-2.8, 0.8] | 0.27 | -1.25 | [-6.1, 3.6] | 0.617 | -0.07 | [-5.0, 4.8] | 0.978 | 0.7 | [-0.8, 2.2] | 0.372 | 4.91 | [1.6, 8.2] | 0.004 | 7.38 | [2.9, 11.9] | 0.001 |
| | $R^2$ | | | 0.089 | | | 0.089 | | | 0.163 | | | 0.161 | | | 0.215 | | | 0.226 |
| Model 6 | B | -0.27 | [-1.0, 0.5] | 0.478 | 0.15 | [-2.0, 2.3] | 0.888 | 0.65 | [-1.5, 2.8] | 0.548 | 0.92 | [0.4, 1.4] | 0.000 | 0.77 | [-0.3, 1.8] | 0.144 | 2.26 | [0.7, 3.8] | 0.004 |
| | C | -0.42 | [-1.2, 0.3] | 0.284 | 1.45 | [-0.7, 3.6] | 0.189 | 2.39 | [0.2, 4.6] | 0.032 | 1.30 | [0.7, 1.9] | 0.000 | 2.10 | [1.0, 3.2] | 0.000 | 4.59 | [3.0, 6.2] | 0.000 |
| | D | 0.95 | [0.0, 1.9] | 0.0407 | 6.80 | [4.2, 9.4] | 0.000 | 7.76 | [5.2, 10.3] | 0.000 | 1.64 | [0.9, 2.4] | 0.000 | 5.58 | [4.1, 7.1] | 0.000 | 9.74 | [7.5, 12.0] | 0.000 |
| | $R^2$ | | | 0.277 | | | 0.275 | | | 0.266 | | | 0.384 | | | 0.489 | | | 0.403 |

Variable Set 2 contained wall thickness data of 16 AHA segments at end-diastole and end-systole, plus ejection fraction, heart rate, end-diastolic and end-systolic volume, and myocardial mass. Beta denotes coefficient from linear regression with exposure remodeling cluster (levels B, C, D, with A as the reference level) and outcome risk score (SCORE2, FRS10, FRS30). Adjustment was as follows: 1) none, 2) end-diastolic mass, 3) average overall end-diastolic wall thickness, 4) concentricity, 5) concentricity and average overall end-diastolic wall thickness, 6) systolic and diastolic blood pressure. $R^2$ denotes percentage of variance in risk scores explained.

Supplementary Table S5: Associations of clusters of remodeling patterns with incident outcome (composite of myocardial infarction, stroke, heart failure and mortality).

| | | Men | | | | | | Women | | | | | |
|---|---|---|---|---|---|---|---|---|---|---|---|---|---|
| | | Composite Outcome | | | Composite CVD Outcome | | | Composite Outcome | | | Composite CVD Outcome | | |
| | | OR | 95% CI | p-value | OR | 95% CI | p-value | OR | 95% CI | p-value | OR | 95% CI | p-value |
| Model 1 | B | 1.54 | [0.91, 2.61] | 0.108 | 1.96 | [1.06, 3.65] | 0.033 | 1.28 | [0.61, 2.65] | 0.515 | 1.84 | [0.7, 4.82] | 0.216 |
| | C | 1.42 | [0.83, 2.42] | 0.198 | 1.40 | [0.73, 2.69] | 0.312 | 2.09 | [1.02, 4.31] | 0.044 | 2.95 | [1.14, 7.63] | 0.026 |
| | D | 2.25 | [1.29, 3.91] | 0.004 | 2.32 | [1.19, 4.51] | 0.013 | 1.61 | [0.61, 4.25] | 0.335 | 2.31 | [0.68, 7.79] | 0.178 |
| Model 2 | B | 1.72 | [0.99, 2.97] | 0.053 | 2.01 | [1.06, 3.82] | 0.033 | 1.43 | [0.67, 3.06] | 0.360 | 1.98 | [0.73, 5.37] | 0.178 |
| | C | 1.71 | [0.95, 3.09] | 0.075 | 1.46 | [0.71, 2.98] | 0.301 | 2.59 | [1.14, 5.92] | 0.024 | 3.41 | [1.18, 9.87] | 0.024 |
| | D | 3.01 | [1.52, 5.97] | 0.002 | 2.47 | [1.10, 5.56] | 0.028 | 2.28 | [0.71, 7.29] | 0.166 | 2.92 | [0.69, 12.29] | 0.144 |
| Model 3 | B | 1.40 | [0.76, 2.60] | 0.279 | 1.83 | [0.89, 3.78] | 0.100 | 0.93 | [0.39, 2.23] | 0.869 | 1.11 | [0.37, 3.39] | 0.851 |
| | C | 1.21 | [0.57, 2.59] | 0.624 | 1.24 | [0.50, 3.09] | 0.639 | 1.15 | [0.36, 3.67] | 0.813 | 1.14 | [0.28, 4.73] | 0.854 |
| | D | 1.74 | [0.63, 4.86] | 0.287 | 1.92 | [0.58, 6.43] | 0.288 | 0.59 | [0.10, 3.65] | 0.573 | 0.47 | [0.05, 4.19] | 0.497 |
| Model 4 | B | 1.35 | [0.77, 2.36] | 0.295 | 1.75 | [0.92, 3.33] | 0.087 | 0.66 | [0.30, 1.45] | 0.298 | 0.97 | [0.35, 2.67] | 0.958 |
| | C | 1.12 | [0.64, 1.98] | 0.689 | 1.12 | [0.57, 2.21] | 0.736 | 0.72 | [0.32, 1.59] | 0.415 | 1.03 | [0.37, 2.85] | 0.950 |
| | D | 1.61 | [0.90, 2.89] | 0.112 | 1.70 | [0.85, 3.37] | 0.132 | 0.52 | [0.18, 1.49] | 0.226 | 0.77 | [0.21, 2.80] | 0.696 |
| Model 5 | B | 1.40 | [0.76, 2.57] | 0.275 | 2.15 | [1.01, 4.59] | 0.048 | 0.91 | [0.41, 2.01] | 0.817 | 1.37 | [0.47, 4.04] | 0.567 |
| | C | 1.06 | [0.56, 2.00] | 0.852 | 1.33 | [0.59, 2.99] | 0.486 | 1.26 | [0.57, 2.82] | 0.568 | 2.30 | [0.79, 6.68] | 0.126 |
| | D | 1.22 | [0.62, 2.41] | 0.569 | 1.74 | [0.75, 4.05] | 0.198 | 0.67 | [0.21, 2.11] | 0.491 | 1.29 | [0.31, 5.31] | 0.728 |

Odds Ratio (OR) from logistic regression with exposure remodeling cluster (levels B, C, D, with A as the reference level). Composite outcome: myocardial infarction, stroke, heart failure, and all-cause mortality. Incident CVD outcome: myocardial infarction, stroke, heart failure, and CVD mortality. Adjustment was as follows: 1) none, 2) end-diastolic mass, 3) average overall end-diastolic wall thickness, 4) age, 5) FRS10.

### Supplementary Figures

Supplementary Figure S1: Participant flowchart

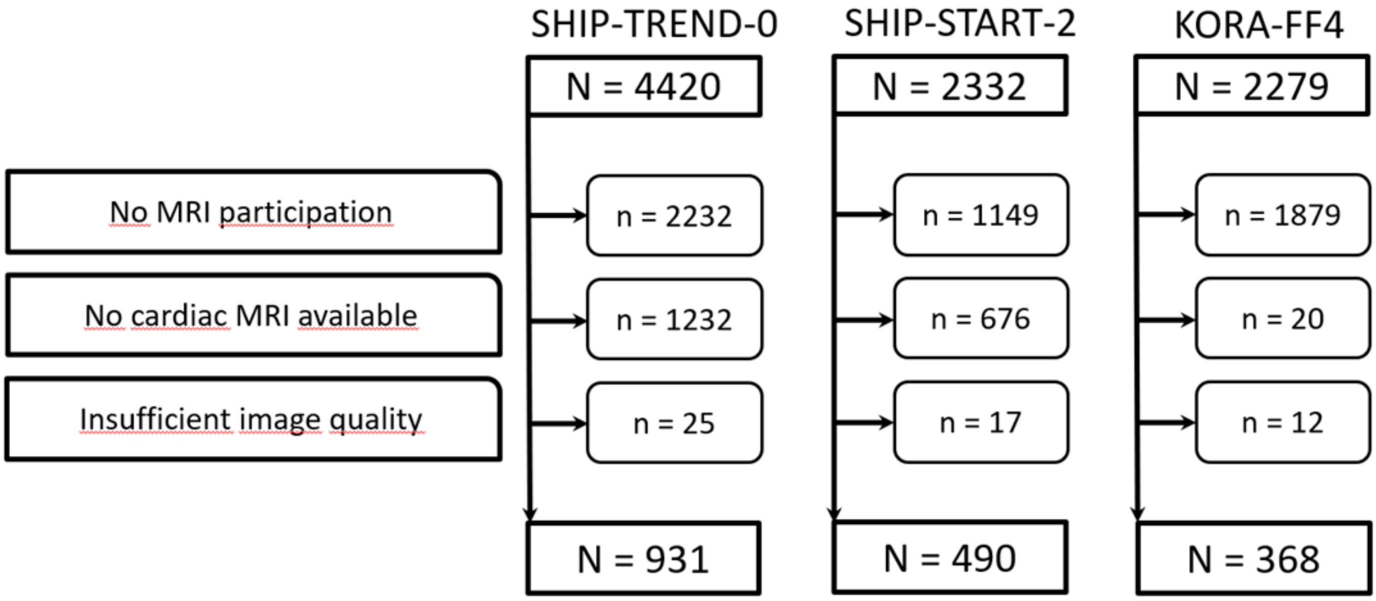

Supplementary Figure S2: Distribution of estimated CVD risk according to quartiles of end-diastolic myocardial mass (upper panel), average overall end-diastolic wall thickness (middle panel), and concentricity (lower panel).

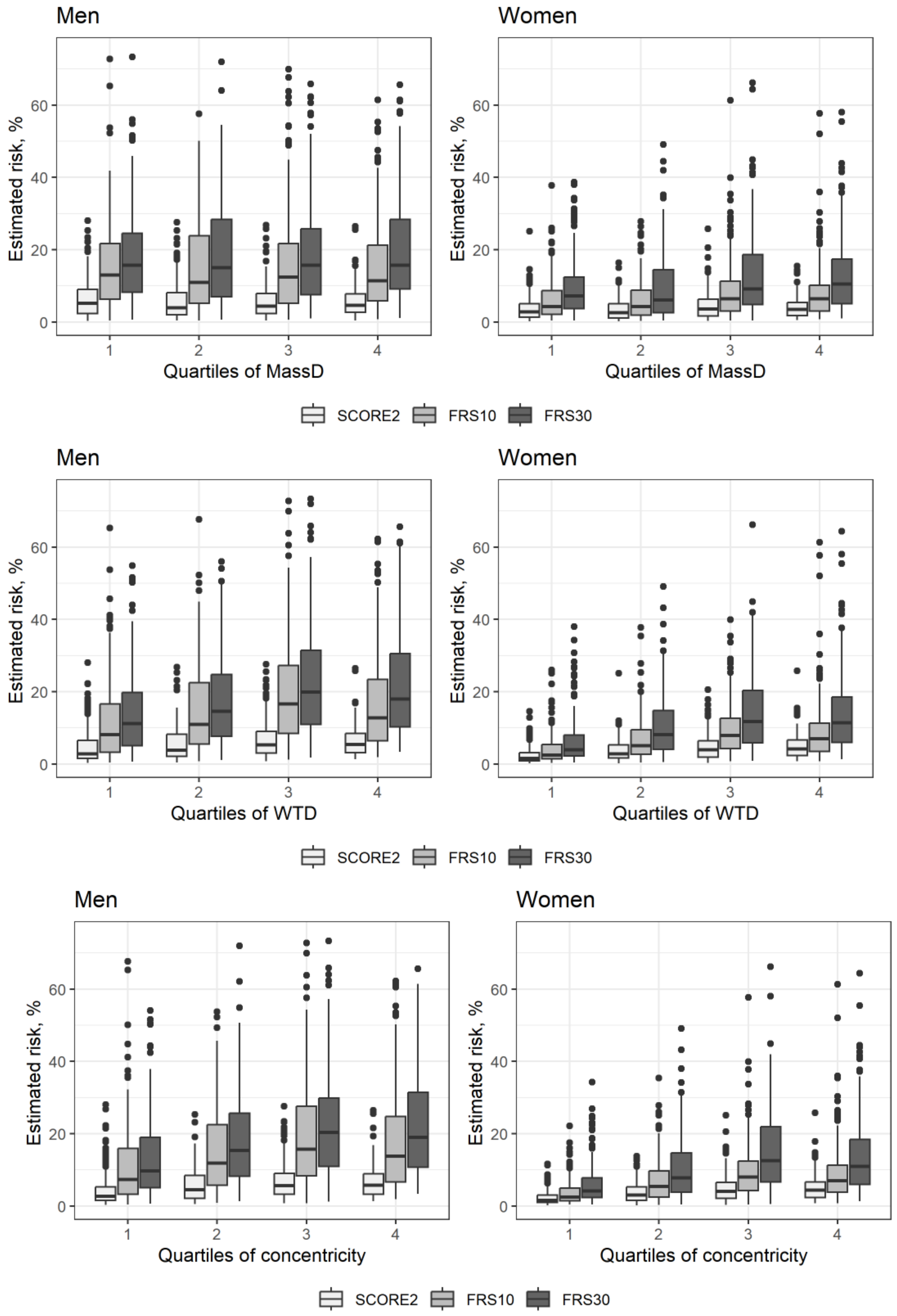

Supplementary Figure S3: Incident outcomes according to clusters. On the x-axis: Cluster, representing phenotypes of LV remodeling patterns. On the y-axis: proportion of total (sex-stratified) sample. Shaded areas denote mortality. Unshaded areas denote composite outcome of incident myocardial infarction, stroke, or heart failure, and mortality.

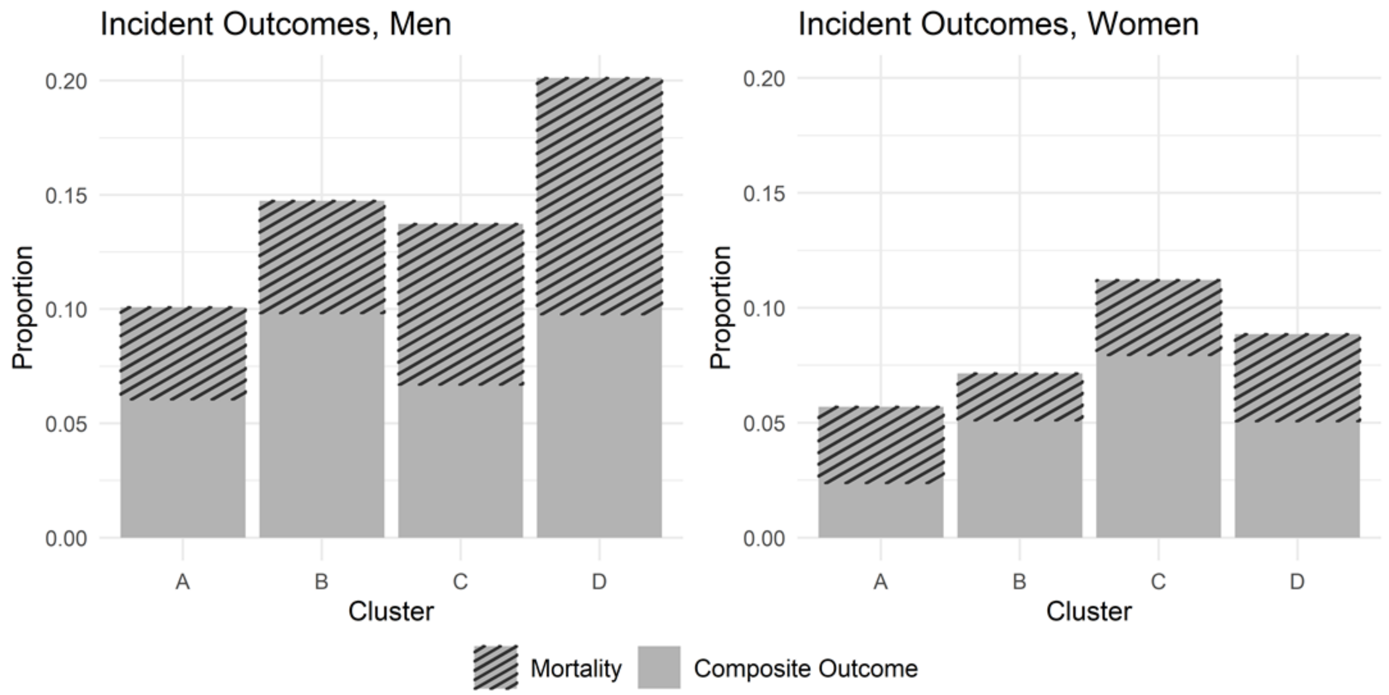
